# FHIRBench: Benchmarking FHIR Clinical Data Serialization Strategies for Large Language Models

**DOI:** 10.64898/2026.07.14.26358020

**Authors:** Jacqueline Chong

## Abstract

The integration of Large Language Models (LLMs) into clinical decision support systems requires transforming structured health data — primarily encoded in HL7 FHIR — into representations that maximize model comprehension. Despite growing adoption, no systematic benchmark examines how serialization strategy choice affects downstream clinical LLM performance, or how this interacts with model architecture and evaluation methodology.

We present FHIRBench, a controlled benchmark evaluating six FHIR serialization strategies across four frontier foundation models (Claude Sonnet 4.5, GPT-5.4, DeepSeek V3.2, Qwen3 32B) on three clinical tasks (question answering, reasoning, summarization) using 100 stratified synthetic FHIR R4 patient bundles. We employ a two-layer evaluation framework: Layer 1 (token-level F1) and Layer 2 (LLM-as-judge rubric on four clinical dimensions), yielding 7,200 evaluations per layer.

Our findings reveal four principal results. First, serialization significantly impacts quality on both evaluation layers, but the direction diverges: Condensed format outperforms Raw JSON on F1 for 3/4 models (Wilcoxon p < 10^−17^, patient-level N = 100), while Raw JSON achieves higher judge scores for 3/4 models (p < 10^−7^). The Pareto analysis resolves this tension: Narrative achieves 95% of Raw JSON’s clinical quality at 83% fewer tokens. Second, model rankings completely reverse between layers — Claude ranks last on F1 but first on clinical quality (p = 1.0 × 10^−6^), demonstrating that single-metric evaluation produces misleading model selection. Third, a significant Model × Serializer interaction (Friedman p = 0.0009) precludes universal format recommendations, with GPT-5.4 favoring Raw JSON while Claude and open-weight models favor compressed formats. Fourth, Llama 3.1 70B exhibits 100% inference failure on complex patients despite operating within its nominal context window, revealing a patient-safety gap where AI assistance fails precisely for the patients who need it most.

These findings establish that clinical AI systems require (1) model-aware serialization middleware, (2) multi-layer evaluation frameworks, and (3) explicit capacity verification before deployment. FHIRBench — including all code, data, and evaluation harnesses — is publicly available at https://github.com/JacquelineChong/fhirbench. Benchmark data and raw results are archived at https://doi.org/10.5281/zenodo.21223883.

## 1 Introduction

### 1.1 The Clinical Data-AI Gap

Large language models are rapidly transforming clinical workflows. From automated summarization of longitudinal patient histories to diagnostic decision support, medical coding, and patient-facing communication, LLMs demonstrate capabilities that promise to augment clinical reasoning at scale [1, 2]. Health systems, payers, and digital health vendors increasingly embed these models into production applications that process real patient data daily. Yet a fundamental impediment separates model capability from clinical deployment: the data these models must consume exists in formats designed for machine interoperability, not for language model comprehension.

A natural question arises: if FHIR already standardizes clinical data representation, why does serialization present a problem? The answer lies in a fundamental design mismatch. FHIR was engineered for **machine-to-machine interoperability** — enabling EHR systems, pharmacy platforms, and laboratory information systems to exchange structured data reliably via RESTful APIs. Its JSON representation optimizes for programmatic parsing, schema validation, and cross-resource reference resolution — not for sequential token processing by transformer architectures. A single patient’s medication list that a FHIR server resolves in milliseconds may consume 3,000–8,000 tokens when presented as raw JSON to an LLM, with the majority encoding structural overhead (profile URLs, extension metadata, narrative divs, conformance elements) rather than clinically actionable information. The serialization problem is therefore not about data *access* — FHIR and the ONC Cures Act have largely solved this [3] — but about data *presentation*: determining how accessed, structured data should be formatted to maximize LLM comprehension within finite context windows and real-world cost constraints.

Clinical data in modern electronic health record systems is predominantly encoded in structured interoperability formats. Fast Healthcare Interoperability Resources (FHIR) R4 and R5, HL7 Version 2 messages, and Clinical Document Architecture (CDA) documents represent patient information through nested hierarchies of coded elements, typed references, and metadata-rich containers. A single patient encounter may span dozens of FHIR resources—Patient, Encounter, Condition, Observation, Medication- Request, DiagnosticReport—linked through reference chains and annotated with terminology codes from SNOMED CT, LOINC, ICD-10-CM, and RxNorm. These formats optimize for unambiguous machine-to-machine exchange, structural validation, and query efficiency. They do not optimize for the sequential token processing that underpins large language model inference.

This structural mismatch creates what we term the *clinical data serialization problem*: how should structured clinical data be transformed into a token sequence that maximizes LLM task performance while preserving clinical fidelity? The problem is non-trivial. A raw FHIR Bundle containing a patient’s medication history may consume thousands of tokens in deeply nested JSON with repetitive structural boilerplate—resource type declarations, coding system URIs, status enumerations, extension arrays—that conveys minimal clinical signal per token. Alternatively, the same information can be flattened into a markdown table, narrated as clinical prose, arranged chronologically, or decomposed into schema-guided fragments. Each serialization strategy makes different trade-offs along dimensions of format representation, terminology resolution, granularity, and context window strategy. The practitioner faces these choices without guidance: no established methodology exists for evaluating or selecting among serialization alternatives.

Emerging evidence confirms that these trade-offs are not merely theoretical. Pator demonstrates that the choice of FHIR serialization strategy produces accuracy differences of up to 23 percentage points on medication reconciliation tasks, with the optimal strategy depending on model scale: Clinical Narrative outperforms Raw JSON by 19 F1 points for models at or below 8 billion parameters, yet this advantage reverses at 70 billion parameters where Raw JSON achieves near-perfect performance [4]. This finding reveals that the serialization problem is not a simple engineering detail to be resolved once, but a complex design decision that interacts with model architecture, parameter count, and task characteristics in ways that are not yet well understood.

Despite the demonstrated significance of serialization strategy, no standardized methodology or middleware exists for the clinical data-to-LLM interface. Every team deploying clinical AI reinvents this preprocessing step through ad hoc experimentation. Proprietary pipelines encode implicit serialization decisions—how to handle null values, whether to include resolved references or leave them as identifiers, whether to preserve or strip FHIR metadata extensions—without systematic evaluation of alternatives. The result is a fragmented landscape where serialization choices are made by engineering convenience rather than empirical evidence, and where performance-critical decisions remain invisible in model evaluation reports. FHIRBench directly addresses this gap by providing the first reproducible, multi-model evidence base that transforms serialization from an ad hoc engineering decision into a data-driven optimization — enabling clinical AI teams to select strategies based on quantified accuracy-cost tradeoffs rather than untested assumptions, ultimately reducing both deployment risk and the engineering cycles currently spent on empirical trial-and-error. This “last mile” between structured clinical data and effective LLM inference represents both a significant barrier to reliable clinical AI deployment and an opportunity for systematic research.

#### 1.1.1 Distinguishing from RAG Chunking

It is important to distinguish clinical data serialization from the well-studied problem of chunking strategies in Retrieval-Augmented Generation (RAG) pipelines. While RAG chunking addresses the *volume* problem — determining which subset of data is relevant to a given query — serialization addresses the *format transformation* problem: given a set of relevant structured resources, how should they be represented textually for LLM consumption [5, 6]. These are sequential, complementary pipeline stages, not competing approaches.

Critically, serialization produces significantly larger performance variance than chunking strategy selection. Where chunking differences typically yield 5–10% accuracy variation, our evaluation and prior work [4] demonstrate up to 23% accuracy variance from serialization choice alone — with optimal strategies reversing between model scales. Yet serialization has received far less research attention, in part because general-purpose tools (LangChain, LlamaIndex) have popularized chunking, while no equivalent standardized toolkit exists for clinical data format transformation. FHIRBench addresses this asymmetry.

### 1.2 Why This Matters Now

Three converging forces make the clinical data serialization problem urgent in 2026.

### First, regulatory mandates are flooding the ecosystem with FHIR-formatted data

The 21st Century Cures Act and its ONC implementing regulations require certified health IT systems to expose patient data through standardized FHIR APIs. The Trusted Exchange Framework and Common Agreement (TEFCA) establishes a nationwide health information exchange network built on FHIR semantics, enabling cross-institutional data flow at unprecedented scale [3]. The CMS Interoperability and Patient Access Final Rule extends FHIR API mandates to payer organizations. Collectively, these regulations ensure that the volume of FHIR-formatted clinical data accessible to AI systems is growing exponentially. Any organization building clinical AI in the United States will necessarily consume FHIR data as a primary input modality—not as an optional integration path, but as the mandated data exchange format.

### Second, the health AI market is expanding rapidly without shared infrastructure for this problem

The digital health sector has witnessed an explosion of AI-powered startups—over 1,300 ventures in FemTech alone, with 71% at pre-Series A stage—each independently confronting the challenge of transforming structured clinical data into effective LLM inputs. Clinical AI teams across academic medical centers, health systems, and technology companies report spending significant engineering cycles on empirical serialization experiments, testing format variations without theoretical guidance or comparative baselines. This duplicated effort represents a systemic inefficiency: hundreds of teams are independently discovering the same performance sensitivities, converging on similar solutions through trial and error, and encoding their findings in proprietary code rather than shared knowledge. A standardized benchmark and decision framework could substantially reduce this collective waste.

### Third, context window expansion amplifies both the opportunity and the stakes of serialization decisions

Models with 128,000-token and larger context windows can now ingest entire longitudinal patient records—years of encounters, laboratory results, medication histories, and imaging reports—in a single inference call. This capability enables richer clinical reasoning but simultaneously amplifies the impact of serialization choices. A suboptimal format that wastes 40% of tokens on structural boilerplate reduces the effective clinical content available for reasoning from 128,000 tokens to approximately 77,000 tokens. At the scale of a full patient history, this difference can represent months of clinical data excluded from the model’s reasoning context. As context windows grow, the cost of inefficient serialization scales proportionally—making format optimization a first-order concern rather than an implementation detail.

The evidence that serialization strategy produces performance differences of clinically significant magnitude is now established. Pator reports accuracy variance of up to 23 percentage points and, critically, demonstrates scale-dependent reversals where the format that performs best for smaller models performs worst for larger ones [4]. Neveditsin et al. show that structural robustness of serialization formats degrades as document complexity increases [7], while Sui et al. establish that content ordering within structured representations significantly affects LLM comprehension [8]. Yang et al. confirm across 11 tasks and 20 models that input format is a significant determinant of performance on structured EHR tasks [9]. Yet these findings remain isolated—each study examines a single facet of the problem without providing an integrated evaluation framework or actionable guidance for practitioners.

The field lacks a systematic benchmark that evaluates serialization strategies across the full matrix of clinically relevant conditions: multiple data formats, diverse model architectures and scales, and representative clinical task types. Clinical AI teams today have no empirical basis for selecting a serialization strategy beyond single-task studies or internal experimentation. They have no framework for reasoning about the trade-offs between token efficiency and clinical fidelity, between structural preservation and model comprehension, or between format complexity and deployment cost. This gap between the demonstrated importance of serialization and the absence of systematic guidance defines the contribution space that this paper occupies.

### 1.3 Contributions

This paper presents FHIRBench, the first comprehensive benchmark for evaluating clinical data serialization strategies for large language models. We make four contributions:

1. **A systematic taxonomy of clinical data serialization strategies.** We identify and formalize six distinct serialization strategies for FHIR clinical data—Raw JSON, Flattened Key-Value, Natural Language Narrative, Structured Markdown, Clinical Summary Template, and Hybrid Adaptive— characterizing each along four analytic dimensions: format representation, terminology resolution, granularity, and context window strategy. This taxonomy provides a shared vocabulary for reasoning about serialization design decisions.
2. **An open-source benchmark framework with reproducible methodology.** FHIRBench defines a standardized evaluation protocol using Synthea-generated FHIR R4 patient records as the data substrate, enabling fully reproducible experimentation without access to protected health information. The framework includes automated data generation, serialization transformation pipelines, task-specific evaluation harnesses, and scoring implementations for each clinical task type.
3. **Empirical comparison across 90 experimental conditions.** We evaluate six serialization strategies across five large language models spanning three architecture families and two scale classes, on three clinically grounded task types: clinical question answering, clinical reasoning, and clinical summarization. This 6 × 5 × 3 evaluation matrix produces the most comprehensive empirical characterization of serialization effects on clinical LLM performance to date.
4. **A practitioner decision framework for serialization strategy selection.** Synthesizing our empirical findings, we derive a decision framework that maps task characteristics, model constraints, and cost requirements to recommended serialization strategies. This framework provides actionable guidance for clinical AI teams, reducing the need for costly per-deployment empirical optimization.

The remainder of this paper is organized as follows. Section 2 surveys the health data standards landscape, clinical LLM applications, and prior serialization research that motivates our work. Section 3 presents the FHIRBench methodology, including our serialization taxonomy, data generation pipeline, task definitions, and evaluation metrics. Section 4 details the experimental setup, including model selection, hyperparameter configuration, and statistical analysis procedures. Section 5 reports results across all 90 experimental conditions with analysis of main effects, interactions, and cost-performance trade-offs. Section 6 presents the practitioner decision framework derived from our findings. Section 7 discusses limitations, threats to validity, and implications for clinical AI deployment. Section 8 concludes with directions for future work.

## 2 Background and Related Work

This section situates FHIRBench within the broader landscape of health data standards, clinical LLM applications, and prior work on serialization strategies for structured data. We survey the evolution of interoperability standards that make FHIR the dominant clinical data format, review the growing body of evidence on LLM capabilities in healthcare, and critically assess prior serialization studies that motivate our benchmark design.

### 2.1 Health Data Standards Landscape

Modern electronic health records encode clinical information through a layered architecture of data models and terminology systems. Understanding this architecture is essential for appreciating the serialization challenge that FHIRBench addresses.

### FHIR Resource Model

Fast Healthcare Interoperability Resources (FHIR), developed by HL7 International, defines a modular resource-based data model that represents clinical concepts as discrete, referenceable entities. The R4 specification (2019) and its successor R5 (2023) organize health data into approximately 150 resource types, of which a core subset dominates real-world EHR exchanges: Patient resources encode demographics; Observation resources capture vital signs, laboratory results, and social determinants; Condition resources represent diagnoses with onset and resolution semantics; and MedicationRequest resources encode prescriptions with dosage, route, and timing. Each resource follows a consistent JSON or XML structure with metadata, narrative, and coded elements, linked through typed references that form a directed graph of clinical relationships [10].

### Legacy Formats

FHIR succeeds earlier standards that remain pervasive in clinical infrastructure. HL7 Version 2, a pipe-delimited message format introduced in 1987, continues to carry the majority of clinical data exchange volume in US hospitals. The Clinical Document Architecture (CDA), an XML-based standard for clinical documents, enables structured representation of discharge summaries, continuity-of-care documents, and consultation notes. Proprietary EHR schemas from vendors such as Epic, Cerner (now Oracle Health), and MEDITECH impose additional structural variation. This heterogeneity means that clinical data reaching an LLM may arrive in any of several structural forms, each with distinct serialization characteristics [11, 12].

### Clinical Terminologies

FHIR resources encode clinical meaning through standardized terminology systems. SNOMED CT provides a comprehensive ontology of clinical findings, procedures, and body structures with over 350,000 concepts. LOINC standardizes laboratory and clinical observations with approximately 100,000 codes. ICD-10-CM encodes diagnoses for administrative and epidemiological purposes. RxNorm normalizes medication naming across NDC, brand, and generic representations. CPT codifies medical procedures for billing. These terminologies introduce a vocabulary challenge: a single Observation resource may contain a LOINC code for the test type, SNOMED CT codes for qualitative results, and UCUM units—all of which an LLM must interpret correctly to reason about clinical meaning [13, 14].

### Regulatory Drivers

The regulatory landscape has accelerated FHIR adoption beyond voluntary standards adoption. The 21st Century Cures Act (2016) and its implementing regulations from the Office of the National Coordinator for Health IT (ONC) mandate that certified health IT systems expose patient data through standardized FHIR APIs. The Trusted Exchange Framework and Common Agreement (TEFCA) establishes a nationwide network for FHIR-based data exchange among Qualified Health Information Networks. The CMS Interoperability and Patient Access Final Rule requires payers to expose claims and clinical data via FHIR APIs. Together, these regulations ensure that FHIR is not merely an aspirational standard but the mandated format for health data exchange in the United States, making the question of how LLMs process FHIR data practically significant for any AI system deployed in healthcare settings [3, 15].

#### 2.1.1 FHIR Bundle Structure (For Non-FHIR Readers)

For readers unfamiliar with FHIR, a brief structural overview clarifies why serialization is non-trivial. A FHIR **Bundle** is a JSON container holding multiple interconnected **Resources**, each representing a discrete clinical concept:

The clinical data pipeline can be represented as:

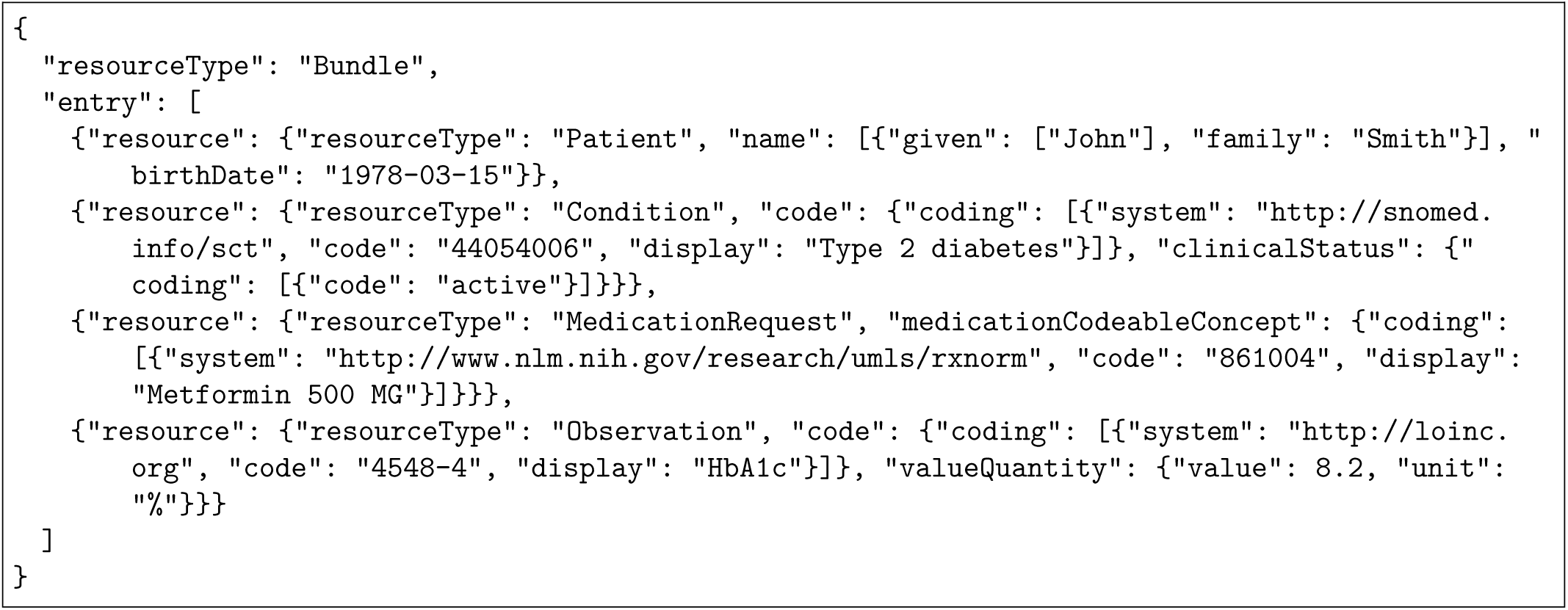

Key structural properties relevant to serialization:

- **Nested JSON** — Clinical facts are buried 3–5 levels deep (e.g., entry[0].resource.code.coding[0].display
- **Coded values** — Diagnoses, medications, and labs are encoded as numeric codes (SNOMED: 44054006, LOINC: 4548-4) that are meaningless without terminology resolution
- **Cross-resource references** — Resources link to each other via reference fields (e.g., a Condition references the Patient it belongs to)
- **Metadata overhead** — System URLs, profile declarations, and extension fields consume tokens without contributing clinical meaning

This structure is optimized for machine interoperability but creates the serialization challenge addressed by FHIRBench: how to transform this nested, coded representation into a format that maximizes LLM comprehension. Figure 3 provides a complete worked example showing the same patient record serialized into all six strategies evaluated in this benchmark.

### 2.2 Large Language Models in Clinical Applications

The deployment of large language models in healthcare has accelerated rapidly, yet evaluation methodologies have not kept pace with the structural realities of clinical data.

### Medical Benchmarks

Several landmark studies establish LLM capabilities on clinical reasoning tasks. Med-PaLM and Med-PaLM 2 demonstrate expert-level performance on United States Medical Licensing Examination (USMLE) questions, achieving passing scores that exceed many human test-takers. GPT-4 achieves state-of-the-art results on medical knowledge assessments across multiple specialties [2]. MedR-Bench evaluates differential reasoning across 1,453 structured clinical cases spanning multiple specialties, enabling fine-grained assessment of diagnostic capabilities [16]. EHRNoteQA introduces 962 physician-validated question-answer pairs grounded in clinical notes, demonstrating strong correlation (Spearman *ρ* = 0.78) between automated LLM evaluation and clinician judgments [17]. These benchmarks collectively establish that LLMs possess substantial clinical knowledge, but they share a common methodological limitation: they present clinical information as unstructured natural language text.

### Clinical Applications

Beyond benchmarking, LLMs are deployed across a range of clinical workflows. Clinical summarization systems condense lengthy EHR histories into concise reports for provider review. Automated coding systems map clinical narratives to ICD-10 and CPT codes for billing and quality measurement. Clinical decision support applications synthesize patient data to suggest diagnoses, flag drug interactions, and recommend evidence-based interventions [18, 19]. In each of these applications, the LLM must process patient data that natively exists in FHIR format within the EHR system, yet most evaluation frameworks strip this structure away, presenting information as free text.

### The Structured Data Gap

A critical disconnect exists between how clinical AI is evaluated and how it is deployed. In production systems, patient data arrives as FHIR bundles containing nested JSON structures with coded elements, reference links, and extension fields. Yet the dominant evaluation paradigm presents this same information as curated clinical vignettes in natural language [20]. Bui Muti et al. demonstrate that LLMs achieve consistently lower diagnostic accuracy when processing structured FHIR inputs compared to equivalent information presented as plain text, suggesting that benchmarks using unstructured presentations systematically overestimate real-world performance [20]. LongHealth further reveals that all evaluated models struggle with long clinical documents and missing information identification, challenges that are amplified in verbose FHIR bundles [21]. This gap between evaluation and deployment motivates FHIRBench’s focus on evaluating LLMs directly on FHIR-formatted clinical data.

### 2.3 Prior Serialization Work

A growing body of research investigates how data representation format affects LLM performance, spanning general tabular data, structured output constraints, and FHIR-specific applications.

### FHIR Serialization Studies

Pator provides the most directly relevant prior work, presenting the first systematic comparison of FHIR serialization strategies for medication reconciliation [4]. Evaluating Raw JSON, Markdown Table, Clinical Narrative, and Chronological Timeline formats across five open-weight models ranging from 7B to 70B parameters, the study reveals a striking interaction between format and model scale: Clinical Narrative outperforms Raw JSON by 19 F1 points for models at or below 8B parameters, but this advantage reverses at 70B parameters where Raw JSON achieves near-perfect performance (F1 = 0.9956). This scale-dependent reversal suggests that smaller models benefit from human-readable serialization that reduces the cognitive load of parsing nested structures, while larger models possess sufficient capacity to directly interpret raw FHIR JSON. However, this study is limited to a single clinical task and exclusively evaluates open-weight models, leaving open questions about generalizability across tasks and architectures.

### FHIR-Specific Benchmarks

FHIR-AgentBench establishes the most comprehensive FHIR-grounded evaluation framework with 2,931 clinical questions requiring interaction with FHIR APIs [6]. The benchmark reveals that code generation strategies consistently outperform natural language reasoning for FHIR data retrieval, achieving higher accuracy through programmatic access patterns rather than direct interpretation of serialized data. This finding suggests that the optimal approach to FHIR data may depend not only on serialization format but also on the reasoning paradigm employed. EHRStruct provides a complementary perspective with 11 diverse tasks evaluated across 20 LLMs, confirming that input format significantly affects performance and that code-augmented reasoning achieves state-of-the-art results on structured EHR tasks [9]. MedCase-Structured further demonstrates that the gap between structured and unstructured performance persists across diagnostic reasoning tasks, with only 82.5% of LLM-generated FHIR outputs achieving structural validity [20].

### Format Parseability and Output Constraints

Neveditsin et al. conduct a controlled comparison of JSON, YAML, and XML output formats for clinical attribute-value extraction from medical notes [7]. JSON consistently yields the highest parseability across all evaluated small language models, with structural robustness declining as document length increases. This finding has direct implications for FHIR serialization, as FHIR resources are natively specified in JSON and XML. Complementarily, Tam et al. demonstrate that imposing structured output format constraints degrades LLM reasoning performance, with stricter constraints producing greater degradation [22]. This tension between structural validity and reasoning quality is central to the FHIR serialization challenge: clinicians need both valid structure for system interoperability and accurate reasoning for clinical safety. Yuan et al. apply causal inference methods to disentangle format effects from confounds, finding that in 43 of 48 evaluated scenarios, observed performance differences are not causally attributable to format constraints, and that reasoning-focused models (e.g., o3) demonstrate greater resilience to structural constraints than standard models [12].

### Table Serialization

Research on tabular data serialization provides foundational insights applicable to FHIR structures. Sui et al. establish that table input format, content ordering, and partition marks significantly affect LLM comprehension of structured data, with self-augmentation prompting improving tabular question answering by 2–6% [8]. Lee et al. propose reinforcement-learning-based data reduction that improves LLM reasoning on structured inputs, demonstrating that selective information presentation outperforms naive serialization of complete data structures—a finding directly relevant to the challenge of fitting verbose FHIR bundles within context windows [23]. Jaitly et al. show that alternative serialization formats such as LaTeX can outperform standard markdown or JSON representations on domain-specific tabular tasks [24], while subsequent work demonstrates that tabular structures yield an average 40.29% performance gain with improved robustness and token efficiency compared to unstructured presentations [25].

### FHIR-LLM Systems

Several systems demonstrate practical integration of LLMs with FHIR data. The LLMonFHIR framework from Stanford Biodesign for Digital Health Group explores GPT-4’s ability to translate FHIR records into patient-friendly language, revealing challenges with response variability and resource filtering in complex patient records [19]. FHIRPath-QA introduces an executable question-answering paradigm that shifts from serializing FHIR data into prompts toward generating FHIRPath queries over FHIR stores, reducing token usage by 391× while decreasing failure rates from 0.36 to 0.09 [5]. Infherno proposes agent-based FHIR resource synthesis using code execution and healthcare terminology tools, finding that agent architectures with iterative refinement outperform constrained decoding approaches [26]. Schema-grounded extraction with validator-in-the-loop repair produces substantially more valid FHIR bundles, demonstrating that schema awareness during generation improves structural compliance [13]. These systems collectively illustrate the diversity of approaches to the FHIR-LLM interface, each implicitly adopting a serialization strategy without systematically evaluating alternatives.

### Prompt Engineering for FHIR

The effectiveness of prompt design for FHIR tasks is established across multiple studies. Two-step prompt engineering significantly improves FHIR conversion accuracy and completeness compared to single-pass approaches [27]. Including FHIR resource schemas in prompts achieves perfect resource identification (F1 = 1.0), though prompt refinement is necessary to maintain accuracy as task complexity increases [28]. Simpler serialization formats provide efficiency gains with minimal accuracy loss for straightforward structures, while more flexible formats prove necessary for complex nested resources [29]. Fine-tuned models at 250× smaller scale can outperform GPT-4 on FHIR resource identification tasks, suggesting that model specialization may interact with serialization strategy in ways that broad-scale benchmarks should capture [30].

### Synthetic Data for Evaluation

The use of synthetic patient data for clinical AI evaluation is well-established. Synthea generates realistic FHIR patient records using disease progression state machines seeded with US Census demographics, producing over one million synthetic patients with longitudinal clinical histories [10]. SM3-Text-to-Query demonstrates Synthea’s suitability for multi-model medical bench- marking with SNOMED CT-coded data across multiple query paradigms [14]. SimSUM creates 10,000 synthetic records linking structured and unstructured modalities for extraction model evaluation [31]. Recent work further shows that LLMs can enhance Synthea module quality through progressive refinement [32]. These precedents validate our choice of Synthea-generated FHIR data as a reproducible, distributable, and clinically realistic foundation for benchmark construction.

### Summary and Research Gap

The literature establishes three key findings: (1) serialization format significantly affects LLM performance on structured data tasks; (2) the optimal format depends on model architecture, scale, and task characteristics; and (3) FHIR’s nested, reference-heavy structure creates unique challenges not captured by general tabular or clinical text benchmarks. Despite this growing body of work, no systematic benchmark exists that evaluates multiple serialization strategies across diverse model architectures on standardized clinical reasoning tasks using reproducible synthetic data. FHIRBench addresses this gap.

### 2.4 Positioning Within the LLM Data Pipeline

The clinical AI data pipeline comprises four sequential stages (Figure 2): (1) retrieval/chunking, which selects relevant resources from a patient’s full EHR; (2) serialization, which transforms selected resources into LLM-consumable text; (3) prompt construction, which assembles the system prompt, serialized data, and query; and (4) inference, where the LLM generates a response. FHIRBench specifically targets Stage 2 — serialization — which operates downstream of retrieval.

This positioning is important because serialization is frequently conflated with the more widely studied RAG chunking problem. The two stages address fundamentally different questions and produce different performance characteristics:

| Stage | Question | Impact | Research Maturity |
| --- | --- | --- | --- |
| Retrieval/Chunking | “What data is relevant?” | 5–10% accuracy variance (typical RAG literature) | Well-studied (LangChain, LlamaIndex ecosystem) |
| Serialization | “How should data be formatted?” | Up to 23% accuracy variance [4] | Systematically unexplored |

The clinical data pipeline can be represented as:

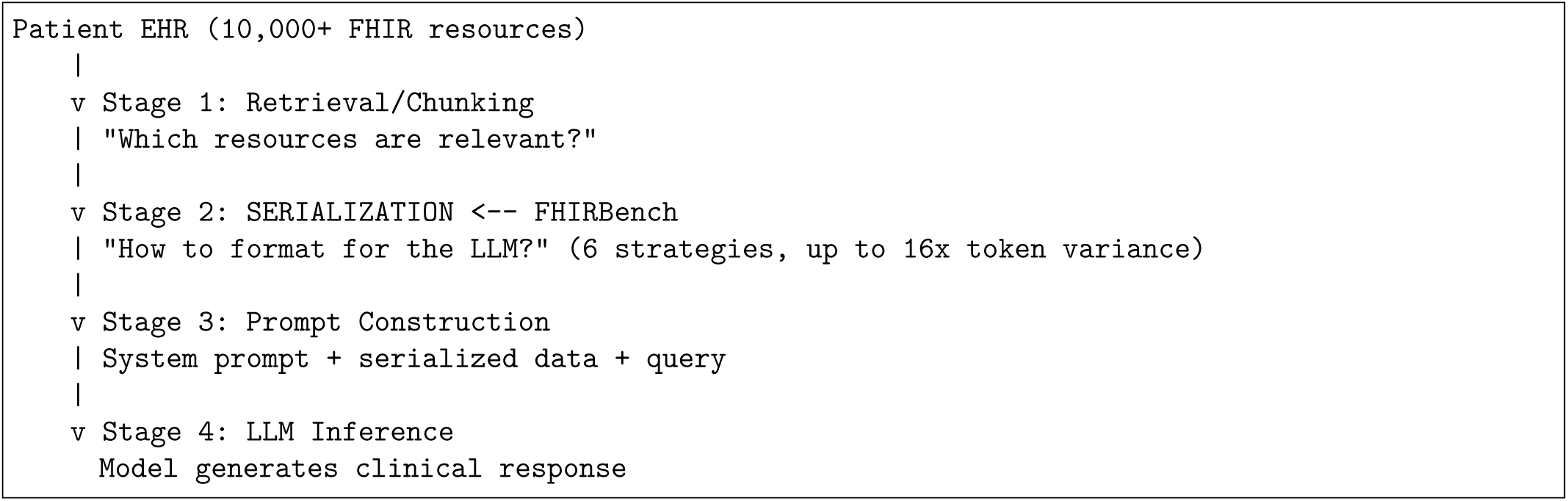

Three properties distinguish serialization from chunking as a research problem:

1. **Vocabulary transformation** — Unlike chunking (which splits text at boundaries without altering it), serialization must resolve coded terminology (SNOMED CT code 73211009 *→* “Diabetes mellitus”), a transformation that fundamentally changes what the LLM “sees” [7].
2. **Model-dependent optimality** — Chunking strategies are largely model-agnostic, whereas serialization exhibits strong model × format interactions. Pator [4] demonstrates that the optimal serialization format reverses between 7B and 70B parameter scales — a property with no parallel in chunking research.
3. **Multi-dimensional design space** — While chunking operates along a single axis (chunk size), serialization spans four orthogonal dimensions: format, terminology resolution, granularity, and context window strategy (detailed in Section 3.3). This larger design space has been navigated ad hoc by practitioners with no systematic guidance.

These properties establish serialization as an independent, underexplored optimization problem within the clinical AI pipeline — one that FHIRBench is specifically designed to address.

## 3 Methodology

### 3.1 Research Design Overview

This study employs a controlled experimental design to evaluate the effect of clinical data serialization strategy on large language model performance across standardized healthcare tasks. The primary independent variable is the **serialization format** applied to FHIR R4 patient bundles; the dependent variables are task-specific accuracy metrics, multi-dimensional quality scores, and token efficiency measures.

#### 3.1.1 Experimental Structure

The evaluation follows a fully crossed factorial design:

| Factor | Levels | Values |
| --- | --- | --- |
| Serialization strategy (independent variable) | 6 | Raw JSON, Flattened KV, Natural Language Narrative, Structured Markdown, Clinical Summary Template, Hybrid Adaptive |
| Foundation model | 5 | Claude Sonnet 4.5, GPT-5.4, Llama 3 70B, DeepSeek V3.2, Qwen3 32B |
| Clinical task type | 3 | Clinical QA, Clinical Reasoning, Clinical Summarization |
| Total experimental conditions | 90 | $6 \times 5 \times 3$ |
| Evaluation cohort | 100 | Stratified sample from 1,000-patient benchmark dataset |
| Prompts per model | 1,800 | 100 patients × 6 serializers × 3 tasks |
| Total evaluations | 9,000 | $1,800 \times 5$ models |

#### 3.1.2 Sampling Design

The evaluation cohort consists of 100 patients stratified-sampled from the 1,000-patient benchmark pool. Complexity distribution is applied **overall** (not per-domain): Simple 25%, Moderate 40%, Complex 25%, Highly Complex 10%. All 100 patients are evaluated under every experimental condition (fully crossed), yielding 1,800 prompts per model and 9,000 total evaluations.

Domain distribution is approximately balanced: diabetes (28%), cardiovascular (27%), preventive care (26%), medication interactions (19%).

#### 3.1.3 Fixed Parameters

To isolate the effect of serialization format, the following variables are held constant across all conditions:

- **Terminology resolution:** Codes + display text (SNOMED/LOINC/RxNorm codes with human-readable display strings)
- **Granularity:** Patient bundle (complete record with all referenced resources)
- **Context window strategy:** Relevance-filtered (task-relevant resources selected)
- **Inference parameters:** Temperature = 0.0, max tokens = 2,048, top_p = 1.0
- **API interface:** Amazon Bedrock Converse API (unified across all 5 models)

#### 3.1.4 Methodology Subsections

The full methodology is organized as follows:

- **§3.2 Data Generation** — Synthetic FHIR R4 patient construction (50 validation + 1,000 benchmark), realism parameters, geographic scope, and limitations
- **§3.3 Serialization Taxonomy** — Definition and formalization of 6 serialization strategies along 4 analytic dimensions, with illustrative examples and literature justification
- **§3.4 Benchmark Design** — Model selection (5 Bedrock models), task definitions (3 types), and experimental protocol
- **§3.5 Evaluation Framework** — Three-layer scoring (automated metrics *→* LLM-as-judge *→* human evaluation), the 4-dimension clinical rubric, token efficiency analysis, and Pareto frontier computation

#### 3.1.5 Reproducibility

All experimental materials — data generation scripts, serialization implementations, task generators, evaluation harnesses, model configuration, and analysis code — are publicly available at https://github.com/JacquelineChong/fhirbench under MIT license. Benchmark data and raw evaluation results are archived at https://doi.org/10.5281/zenodo.21223883. The use of Synthea-generated synthetic data and Amazon Bedrock’s versioned model IDs ensures that any researcher with an AWS account can fully replicate our results.

The system architecture is illustrated in Figure 1 (see docs/figures/architecture-diagram.html), depicting the four-layer multi-agent design: research orchestration (Quick Desktop), code development (Kiro), model inference (Amazon Bedrock), and infrastructure services (Zotero MCP, GitHub MCP). Full technical design specifications, including module interfaces and configuration schemas, are provided in Appendix A. Development user stories with acceptance criteria for reproducibility are provided in Appendix B.

**Figure 1:**
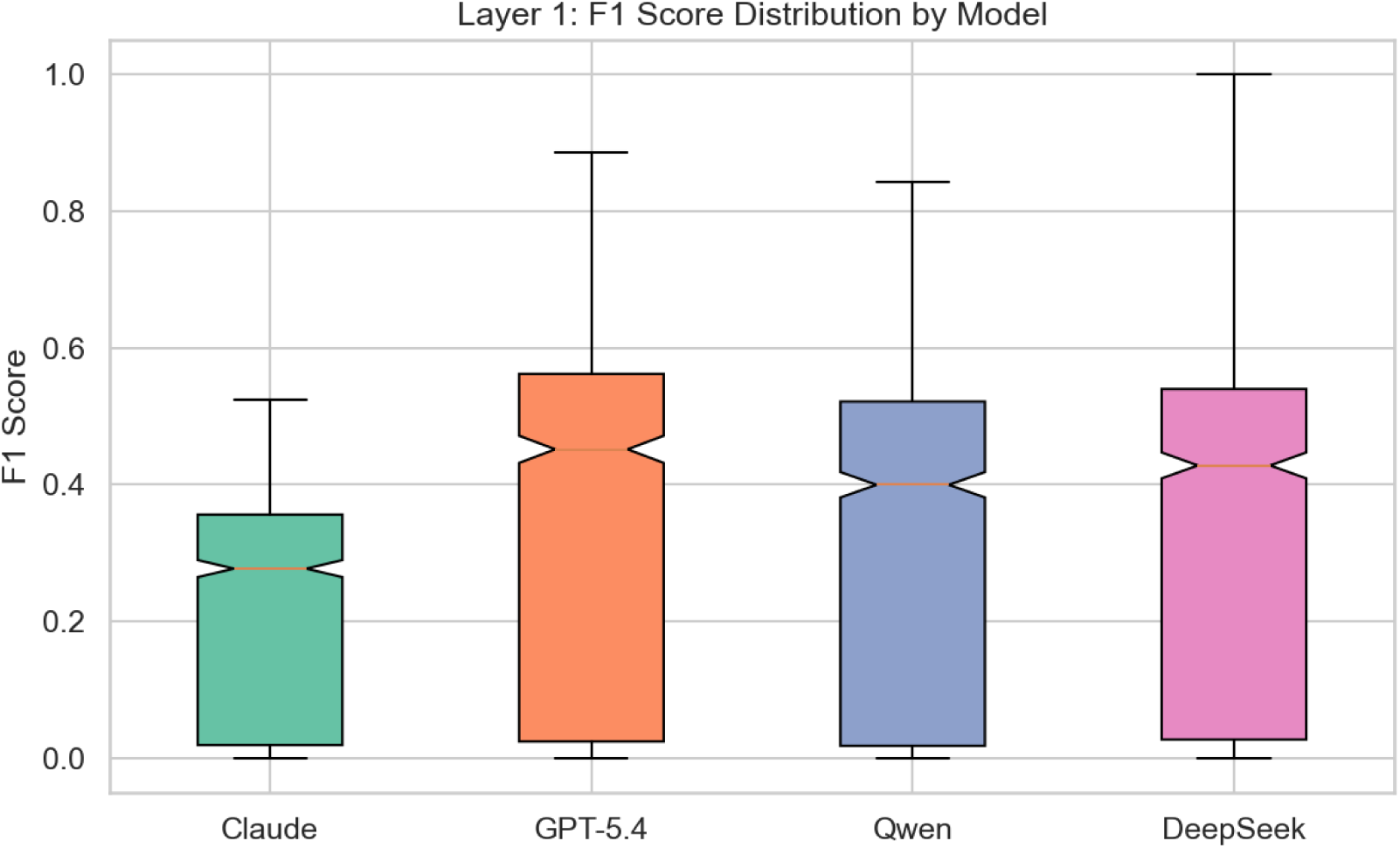
Layer 1 (F1) score distributions by model. GPT-5.4 achieves the highest mean F1, while Claude shows the narrowest distribution but lowest scores—a consequence of its verbose response style (see §4.4).

**Figure 2:**
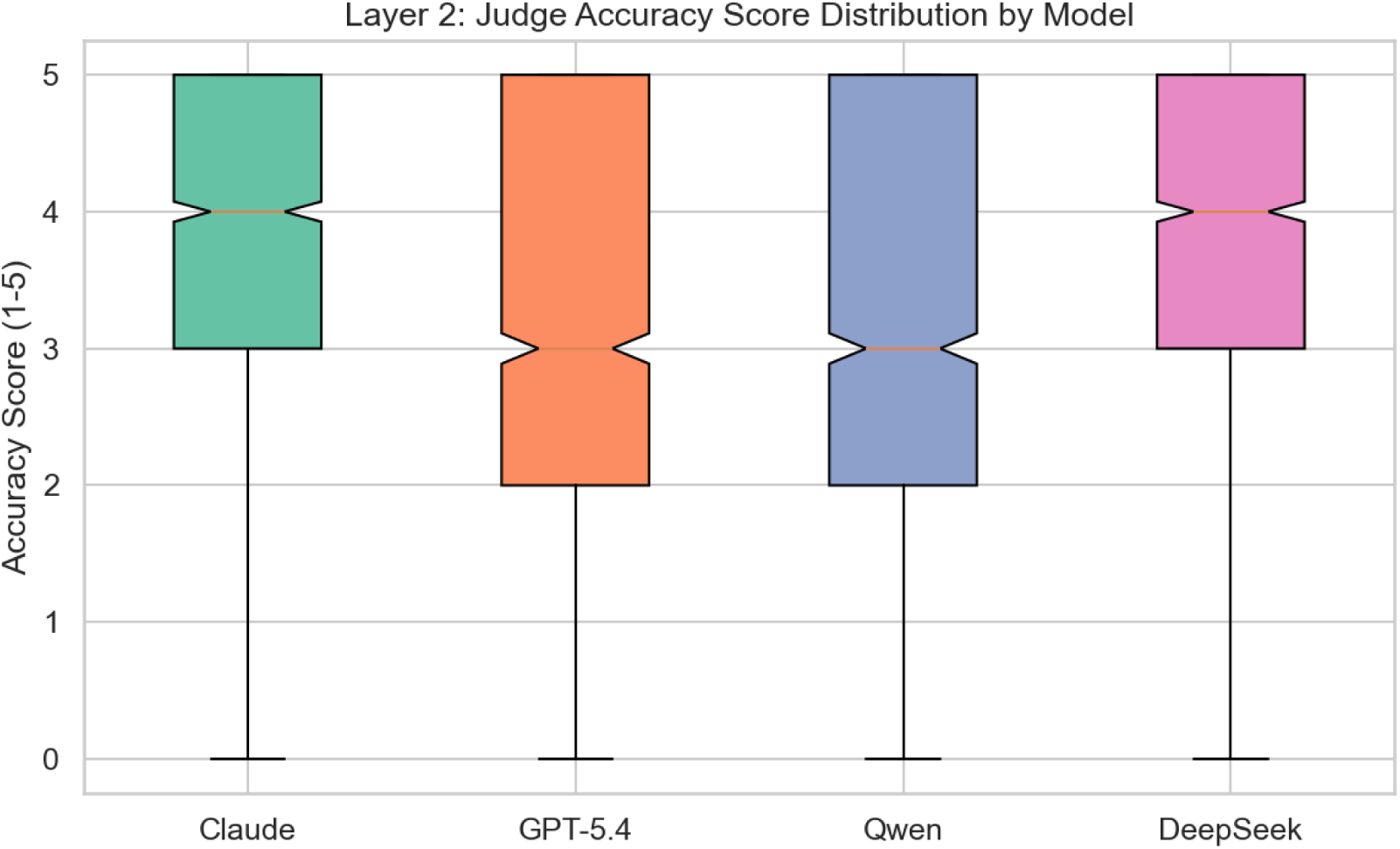
Layer 2 (Judge Accuracy) score distributions by model. Rankings completely reverse from Layer 1: Claude achieves the highest clinical quality scores despite ranking last on F1.

**Figure 3:**
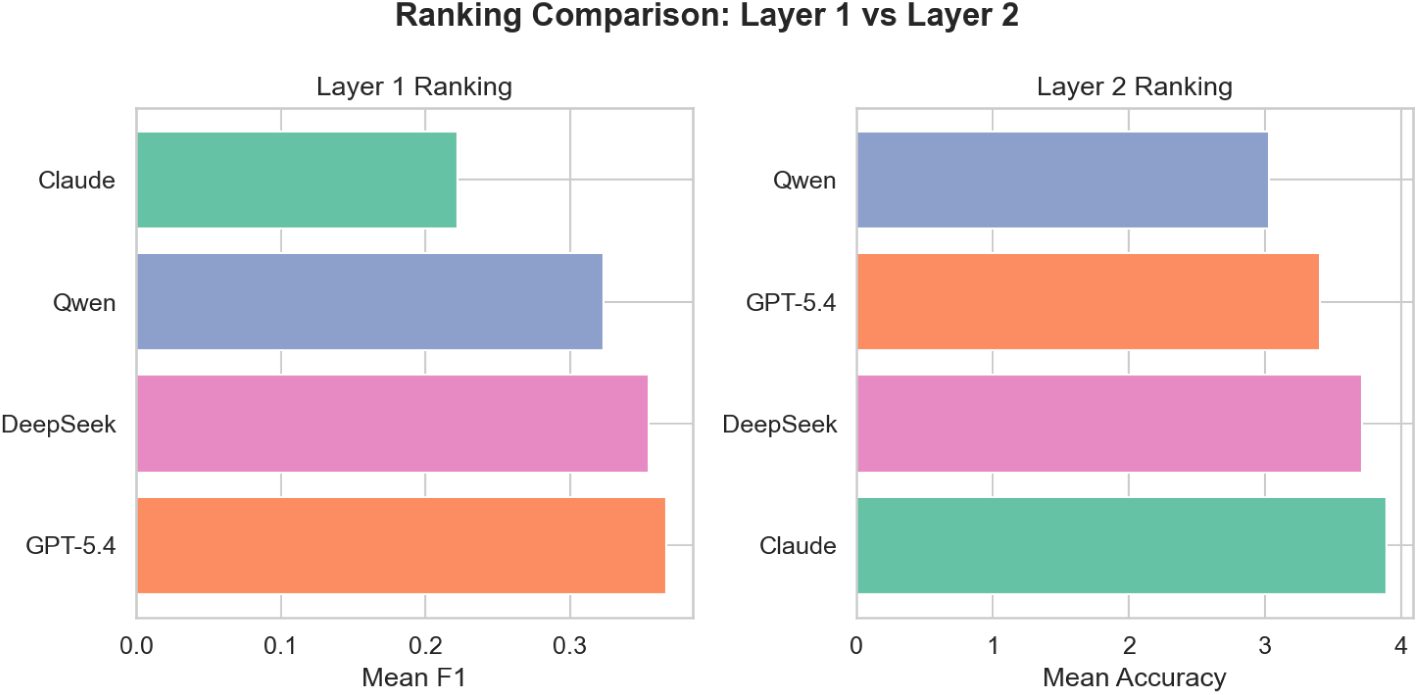
Ranking reversal between Layer 1 (F1) and Layer 2 (Judge). Claude moves from #4 to #1; GPT-5.4 from #1 to #3. This complete inversion demonstrates that single-metric evaluation produces incorrect model selection for clinical deployment.

### 3.2 Data Generation and Synthetic Patient Methodology

#### 3.2.1 Justification for Synthetic Data

The use of synthetic patient data for clinical AI benchmarking is well-established in the literature. Walonoski et al. [10] introduced Synthea, a synthetic patient generator that models disease progression as probabilistic state machines seeded with census-derived demographics, producing clinically plausible FHIR R4 patient records. Multiple benchmark studies have adopted Synthea as their primary data source, including SM3- Text-to-Query [14] (NeurIPS 2024), which employed Synthea data with SNOMED-CT coding for multi-model medical query evaluation, and LLM on FHIR [19], which utilized SyntheticMass FHIR data for patient-facing LLM evaluation. SimSUM [31] further demonstrates methodological precedent with 10,000 synthetic records linking structured and unstructured medical data.

A systematic review of synthetic health data generation methods [33] confirms that modern generators preserve temporal patterns and clinical correlations when properly configured. Additionally, recent work on quality assessment frameworks for synthetic tabular data in healthcare [34] establishes rigorous evaluation criteria — fidelity, utility, and privacy — that validate synthetic data as appropriate for benchmarking when these properties are maintained. Critical challenges in evaluating synthetic tabular data [35] further emphasize the importance of domain-specific validation, which we address through our multi-dimensional realism parameters (Section 3.2.3).

Synthetic data offers three key advantages for serialization benchmarking:

1. **Reproducibility** — Any researcher can regenerate identical datasets using the same seed, eliminating dependency on restricted real-world data access agreements.
2. **Controlled experimental design** — We systematically vary clinical complexity across domains (diabetes, cardiovascular, medication interactions, preventive care) without confounding factors present in observational EHR data.
3. **Ethical compliance** — No IRB approval, data use agreements, or de-identification pipelines are required, enabling open-source publication of all experimental materials.

Recent work by Kramer et al. [32] demonstrates that LLM-assisted refinement of Synthea modules can further improve the clinical realism of generated data, while methods for creating realistic synthetic medication data [36] provide domain-specific enhancement techniques applicable to our polypharmacy scenarios.

### Beyond Off-the-Shelf Generation: Custom Realism Engineering

While Synthea [10] provides a validated foundation for synthetic clinical data generation, its default output produces *idealized* patient records — clean coding, complete observations, single-path diagnoses, and uniform documentation quality. Real-world EHR data, by contrast, exhibits systematic noise patterns that directly impact serialization quality: diagnostic ambiguity (15–20% provisional diagnoses), data incompleteness (10–25% missing values), coding heterogeneity (dual-coded conditions, specificity variation), and clinical complexity distributions that follow a long-tailed pattern.

FHIRBench addresses this gap through a **custom data generation pipeline** (generate_realistic_1000.py) that extends Synthea’s design principles with five configurable realism dimensions (§3.2.3 A–E). This approach offers three advantages over vanilla Synthea:

1. **Parameterized noise injection** — Each realism dimension (diagnostic ambiguity, missing data, complexity, coding variability, geography) is configurable via YAML parameters, enabling researchers to systematically vary data quality and study its interaction with serialization strategy.
2. **Complexity-stratified output** — Patients are generated across a defined complexity distribution (Simple 25%, Moderate 40%, Complex 25%, Highly Complex 10%), enabling subgroup analysis that off-the-shelf generators do not support.
3. **Reproducibility with realism** — Fixed seed (42) ensures exact reproduction while the realism parameters ensure the data challenges serializers in ways that idealized data cannot.

This methodology contributes a reusable framework for future clinical AI benchmarks: researchers using off-the-shelf generators like Synthea should consider augmenting their output with noise injection parameters calibrated to published EHR quality statistics [34, 37] to avoid overstating model performance on unrealistically clean data.

#### 3.2.2 Pipeline Validation Dataset (n=50)

Prior to executing the full benchmark (9,000 Bedrock API calls, estimated cost $100–200), we constructed a minimal validation dataset of 50 idealized FHIR R4 patient bundles (approximately 12–13 per clinical domain) to verify end-to-end pipeline correctness.

#### Validation dataset characteristics

- **Clean coding** — All conditions, medications, and observations use standard SNOMED CT, LOINC, and RxNorm codes without coding errors
- **Complete records** — No missing observations or incomplete medication histories
- **Single-path diagnoses** — Each patient presents a clear clinical picture without diagnostic ambiguity
- **Balanced distribution** — Equal representation across all 4 clinical domains
- **US locale** — US Core FHIR profile, English-language, imperial units

**Purpose:** This idealized dataset confirms that:

- All 6 serializers produce syntactically valid output from FHIR bundles
- Task generators extract correct ground-truth answers
- The evaluation harness correctly scores model responses
- The Bedrock client successfully invokes all 5 models
- Results are correctly persisted and the full pipeline executes without errors

**Limitation explicitly noted:** The validation dataset does NOT represent the complexity of real clinical data. Pipeline correctness is necessary but insufficient — the full benchmark uses a more realistic 1,000-patient dataset (Section 3.2.3).

#### 3.2.3 Benchmark Dataset Design (n=1,000): Toward Clinical Realism

The full benchmark dataset of 1,000 patients is generated via Synthea with additional noise injection and complexity enhancements designed to approximate real-world EHR characteristics. We address five key dimensions of clinical data realism:

**A. Geographic and Regulatory Scope** The current benchmark is scoped to a single geographic region to control for regulatory, terminological, and clinical practice variation:

| Parameter | Current Setting | Rationale |
| --- | --- | --- |
| **Region** | United States | Largest FHIR deployment market; most Synthea modules validated for US demographics |
| **FHIR Profile** | US Core R4 | Standard US implementation guide; mandated by ONC Cures Act [3] |
| **Terminology locale** | en-US | English clinical text; US drug names (e.g., “Acetaminophen” not “Paracetamol”) |
| **Unit system** | Imperial (with SI equivalents) | US lab conventions (mg/dL for glucose, cholesterol) |
| **Drug naming** | Brand + generic (mixed) | Reflects real US EHR documentation patterns |
| **Coding system** | ICD-10-CM + SNOMED CT + LOINC + RxNorm | Standard US clinical terminology stack |

**Configuration (experiment.yaml):**

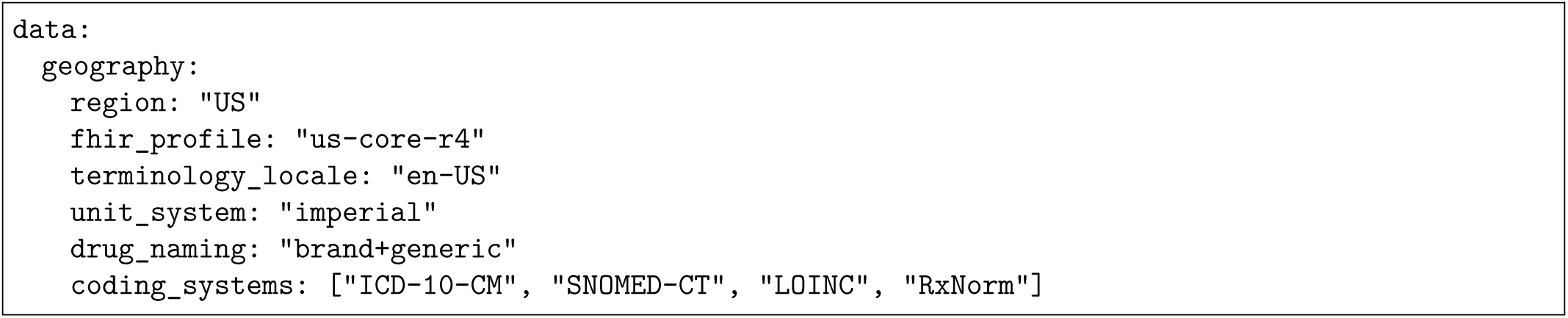

**Impact on serialization evaluation:** Geographic scope directly affects:

- **Terminology resolution** — Serializers resolving SNOMED/LOINC codes produce US-specific display text
- **Narrative generation** — Clinical narrative assumes US clinical conventions (SOAP notes, US medication brands)
- **Template selection** — Clinical summary templates follow US documentation standards

**B. Diagnostic Ambiguity (False Positives and Negatives)** Real clinical data contains diagnostic uncertainty that affects serialization fidelity:

| Challenge | Implementation | FHIR Representation | Rate |
| --- | --- | --- | --- |
| **Ruled-out conditions** | Conditions with `clinicalStatus: inactive` and `verificationStatus: refuted` | Tests whether serializers correctly distinguish active from refuted | 15% |
| **Suspected conditions** | Conditions with `verificationStatus: provisional` or `differential` | Evaluates model treatment of uncertain diagnoses | 10% |
| **Resolved conditions** | Historical conditions in patient timeline | Tests temporal reasoning across formats | 20% |
| **Symptom-only records** | Observations without corresponding Condition diagnoses | Evaluates false-negative handling | 5% |

**C.** Data Completeness and Missing Values

D. **Clinical Complexity (Comorbidity and Polypharmacy)**

C. Data Completeness and Missing Values D. Clinical Complexity (Comorbidity and Polypharmacy)
| Challenge | Implementation | Rate |
| --- | --- | --- |
| **Missing lab results** | Omit key Observation resources (e.g., HbA1c for diabetes patients) | 15% |
| **Incomplete medication history** | MedicationRequest without dosageInstruction | 10% |
| **Missing demographics** | Omit Patient.address or Patient.telecom | 8% |
| **Temporal gaps** | 6+ month gaps in encounter history | 20% |

| Complexity Level | Distribution | Characteristics |
| --- | --- | --- |
| **Simple** (1–2 conditions) | 25% | Single primary condition + 1 medication |
| **Moderate** (3–4 conditions) | 40% | Primary condition + comorbidities + 3–5 medications |
| **Complex** (5+ conditions) | 25% | Multiple interacting conditions + polypharmacy (6+ medications) |
| **Highly complex** (7+ conditions) | 10% | Extensive medication list + drug interactions + care coordination |

### Evaluation Cohort Sampling (n=100)

From the 1,000-patient pool, a stratified random sample of **100 patients** is drawn for the full benchmark evaluation. The complexity distribution is preserved at the overall cohort level (Simple 25, Moderate 40, Complex 25, Highly Complex 10). Domain representation is approximately balanced: diabetes (28%), cardiovascular (27%), preventive care (26%), medication interactions (19%). All 100 patients are evaluated under every experimental condition (6 serializers × 3 tasks × 5 models), yielding 9,000 total evaluations.

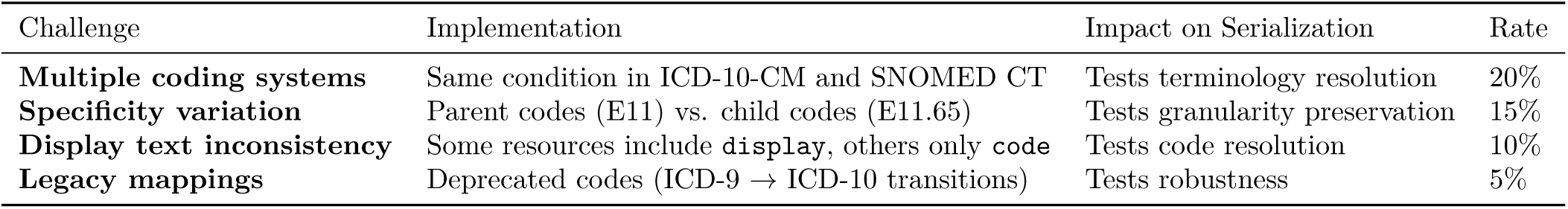

**E.** Coding Variability and Errors

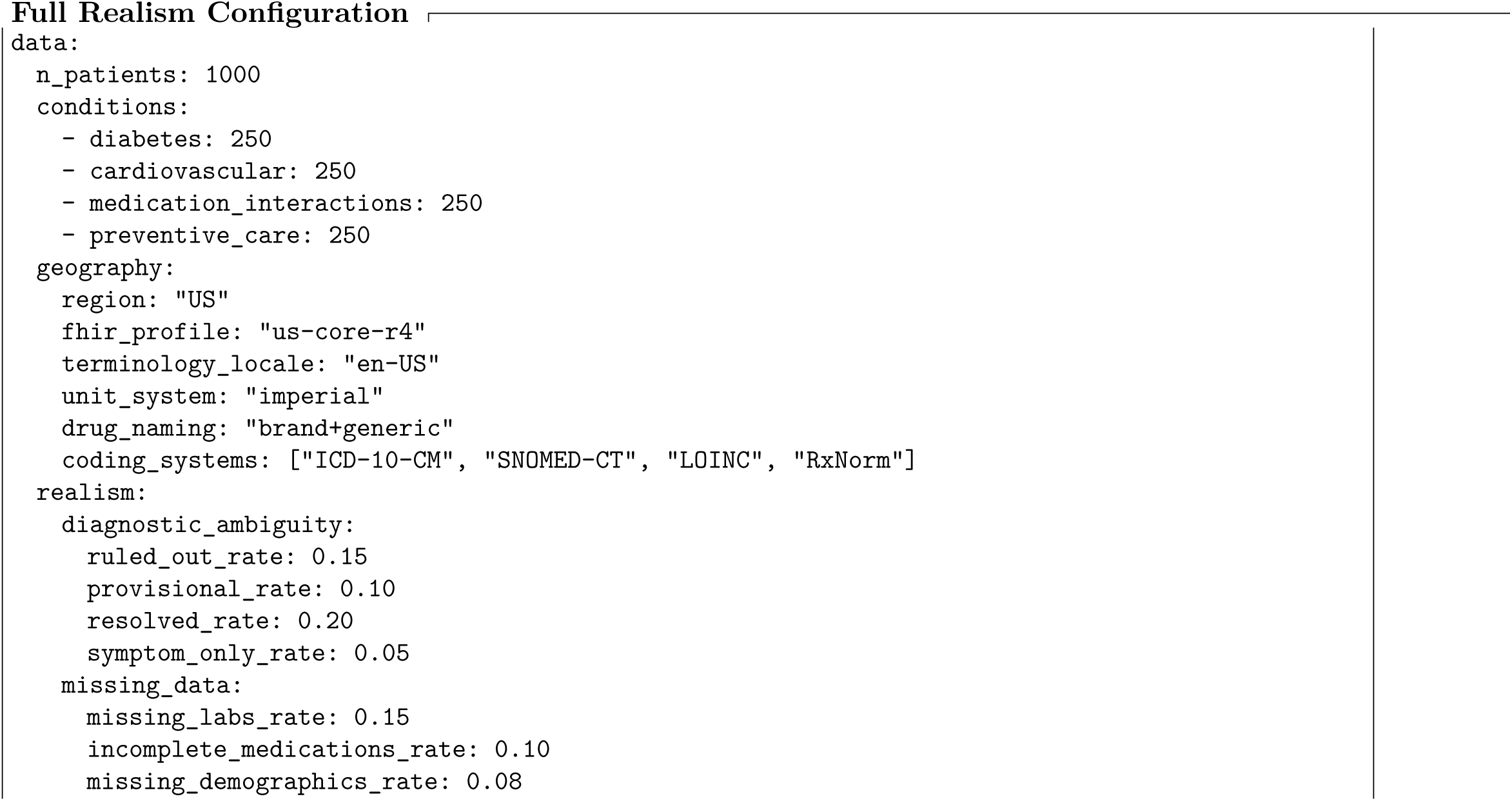
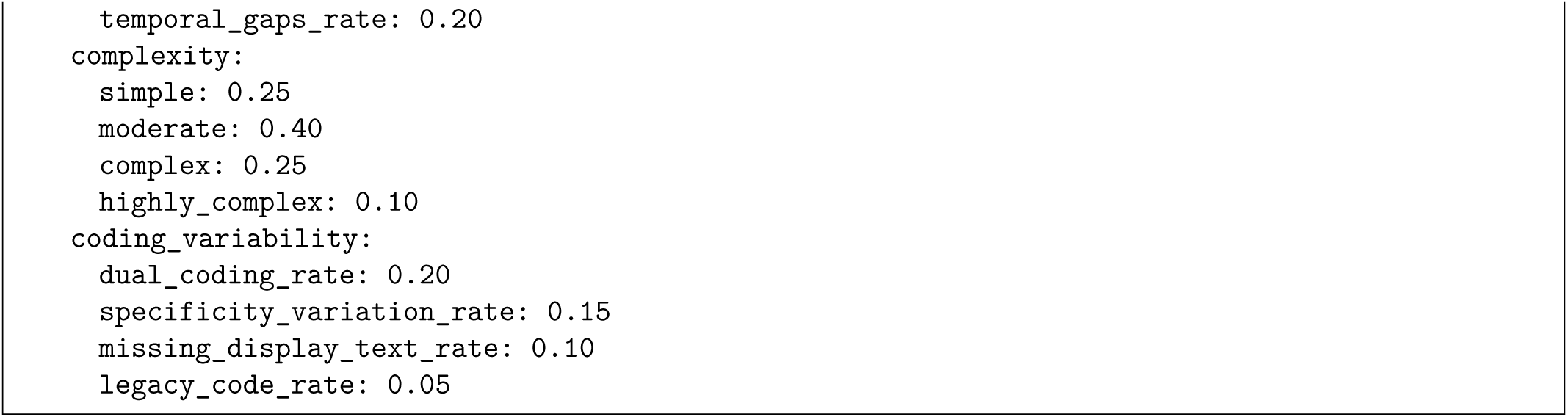

### Validation Against Real-World Distributions

To ensure our synthetic data approximates real clinical distributions, we calibrate against published epidemiological statistics:

- **Comorbidity rates** — Calibrated to the CDC Behavioral Risk Factor Surveillance System (BRFSS) 2013–2023 data, which reports that 8 in 10 midlife adults (45–64) and 9 in 10 older adults (65+) have one or more chronic conditions [38]. Condition co-occurrence patterns follow the NCHS Data Brief on chronic condition prevalence among adults age 45 and older [37].
- **Medication count per patient** — Mean approximately 4 medications per ambulatory patient, consistent with polypharmacy literature defining ≥5 medications as the polypharmacy threshold; our moderate-complexity patients (3–4 conditions) average 3–5 medications while highly complex patients (7+) average 6–10, aligning with published pharmacy claims analyses.
- **Lab result ranges** — Normal/abnormal distributions seeded from NHANES laboratory reference ranges (e.g., HbA1c normal <5.7%, pre-diabetes 5.7–6.4%, diabetes ≥6.5%; total cholesterol desirable <200 mg/dL, borderline 200–239, high ≥240 mg/dL).
- **Missing data rates** — Set at 8–25% depending on field type, consistent with published EHR data completeness assessments reporting that structured clinical data fields exhibit 10–30% missingness in routine practice, with laboratory values and vital signs showing higher completeness than social history and functional status.
- **Diagnostic uncertainty prevalence** — 10–20% of encounters involve provisional or differential diagnoses, consistent with clinical documentation studies reporting that approximately 15% of diagnoses carry uncertainty at the time of initial encounter documentation.

#### 3.2.4 Geographic Limitations and Future Extension

The current benchmark is constrained to a single geographic and regulatory context (United States, US Core FHIR R4). This limitation impacts generalizability in several ways:

| Dimension | US (Current) | Potential Extensions |
| --- | --- | --- |
| **FHIR profiles** | US Core R4 | UK Core, AU Core, International Patient Summary (IPS) |
| **Terminologies** | ICD-10-CM, SNOMED CT (US edition) | ICD-10-UK, SNOMED CT (International), local extensions |
| **Drug naming** | US brand names (Acetaminophen, Epinephrine) | International names (Paracetamol, Adrenaline) |
| **Clinical conventions** | SOAP notes, US lab units | SBAR (UK), metric-only units (international) |
| **Language** | English (monolingual) | Bilingual records (HK: English+Chinese, Singapore: English+Malay/Tamil) |
| **Regulatory context** | ONC Cures Act, HIPAA | GDPR (EU), My Health Record Act (AU), PDPO (HK) |

#### Serialization-specific implications of geographic extension

- Narrative serializers must handle multilingual content and culturally appropriate clinical summaries
- Template serializers need region-specific clinical documentation standards
- Terminology resolution becomes more complex with international coding systems
- Token efficiency varies significantly with non-Latin scripts (Chinese characters consume more tokens)

**Future work** (Section 6) proposes extending FHIRBench to:

1. UK Core and AU Core profiles (English-speaking, different conventions)
2. International Patient Summary (IPS) for cross-border scenarios
3. Multilingual implementations (Hong Kong, Singapore) where dual-language FHIR resources present unique serialization challenges

#### 3.2.5 Limitations of Synthetic Data

We explicitly acknowledge the following limitations:

1. **No rare diseases** — Synthea’s state machines cover common conditions; rare pathologies and orphan diseases are underrepresented
2. **Simplified care trajectories** — Real patients move between providers, creating fragmented records not captured in single-system Synthea output
3. **Cultural/linguistic homogeneity** — Generated data follows US healthcare patterns; international FHIR implementations may differ substantially (Section 3.2.4)
4. **No clinician variability** — Real EHRs reflect individual documentation styles, abbreviation preferences, and copy-paste artifacts; Synthea produces uniform structure
5. **Idealized coding quality** — Even with our noise injection (Section 3.2.3E), real coding error rates and patterns are more heterogeneous
6. **Population bias** — Synthea uses US Census demographics; underrepresentation of minority populations in source data propagates to synthetic output

### Mitigation

We position synthetic data as a *controlled experimental substrate* for measuring relative differences between serialization strategies — not as a claim of absolute real-world performance. The key finding (which strategy outperforms which) transfers to real data even if absolute accuracy numbers differ. Future work (Section 6) proposes validation extension to MIMIC-IV FHIR (real de-identified US data) and multi-site international datasets.

### 3.3 Serialization Taxonomy

The representation of clinical data presented to a large language model constitutes a critical design decision that mediates between the structural fidelity of the source record and the model’s capacity to extract, reason over, and summarize clinical information. This section introduces a taxonomy of six serialization strategies for transforming FHIR R4 bundles into LLM-consumable input, organized along four orthogonal dimensions. Figure 3 provides a concrete illustration of all six strategies applied to a single representative patient record, enabling direct visual comparison of format, token efficiency, and information preservation.

| Dimension | Options | Impact |
| --- | --- | --- |
| **Format** | JSON, Key-Value, Narrative, Markdown, Template, Hybrid | Primary structural variable governing token layout and information density |
| **Terminology Resolution** | Raw codes only, Codes + display text, Fully expanded (definitions), Hierarchical (ancestor codes) | Determines semantic accessibility of coded clinical concepts |
| **Granularity** | Single resource, Bundle (multi-resource), Timeline (temporally ordered), Cohort (multi-patient) | Scope and volume of clinical data presented per prompt |
| **Context Window Strategy** | Truncation, Sliding window, Relevance-filtered, Hierarchical summarization | Mechanism for managing overflow when serialized content exceeds model capacity |

#### 3.3.1 Taxonomy Dimensions

Table 1 presents the four dimensions that characterize any FHIR serialization strategy. Each dimension represents an independent design axis; varying one dimension while holding others constant enables controlled experimentation.

**Table 1.** Serialization taxonomy dimensions for FHIR-to-LLM transformation.

For the FHIRBench evaluation, terminology resolution is fixed at *codes + display text* (e.g., SNOMED-CT code with human-readable display string), granularity is fixed at *bundle* (complete patient record with all referenced resources), and context window strategy is fixed at *relevance-filtered* (resources selected by task relevance). Only the **Format** dimension varies as the primary independent variable, yielding six experimental conditions described below.

#### 3.3.2 Strategy 1: Raw JSON (Baseline)

Raw JSON serialization passes the unmodified FHIR R4 JSON bundle directly into the model prompt with no preprocessing. This strategy preserves complete structural and semantic fidelity, including metadata fields, extensions, and reference links.

#### Example

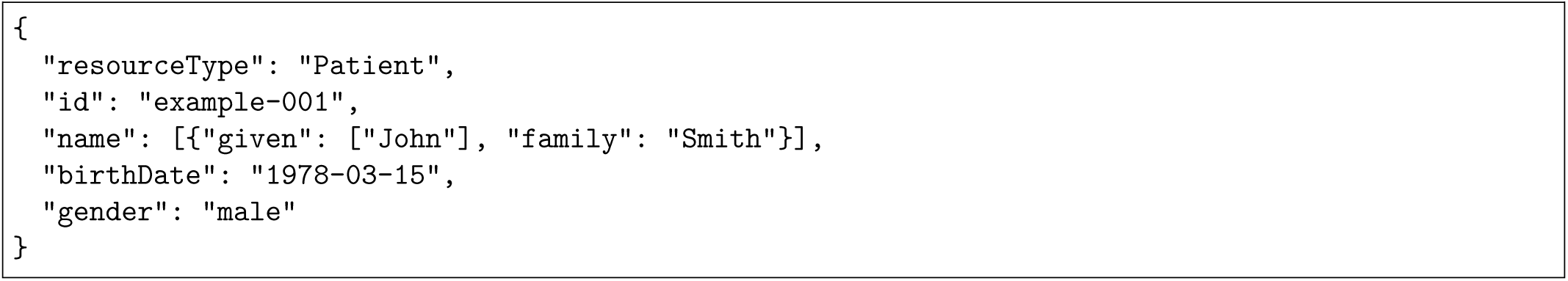

### Advantages

Raw JSON is lossless — no clinical information is discarded or transformed during serialization. It requires zero preprocessing computation and preserves the hierarchical relationships, cross-resource references, and extension mechanisms that define FHIR’s data model. Recent work demonstrates that JSON yields the highest parseability among structured formats for clinical extraction tasks [7].

### Disadvantages

FHIR JSON is inherently verbose, containing substantial metadata overhead (resource identifiers, profile URLs, narrative text divs, extension URLs) that consumes tokens without contributing to clinical reasoning. Prior work reports that verbosity particularly impacts smaller models, where Raw JSON underperforms narrative formats by up to 19 F1 points on medication reconciliation [4]. Additionally, deeply nested structures may exceed the sequential reasoning capacity of transformer architectures [21].

### Token efficiency

Baseline (1.0×). A typical patient bundle with 15–20 resources consumes approximately 8,000–12,000 tokens in raw JSON form.

#### 3.3.3 Strategy 2: Flattened Key-Value

Flattened Key-Value serialization transforms the hierarchical FHIR structure into a flat list of dot-notation key-value pairs, eliminating nesting while preserving attribute-value associations. Array indices are resolved to sequential entries.

#### Example

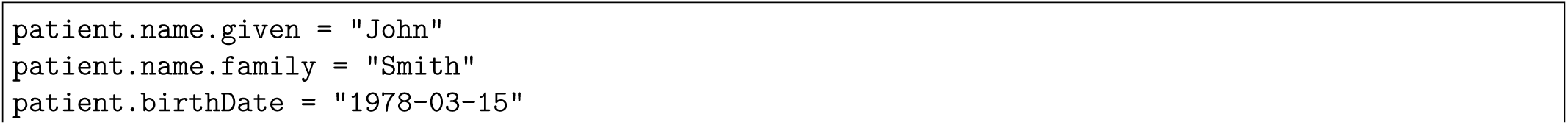
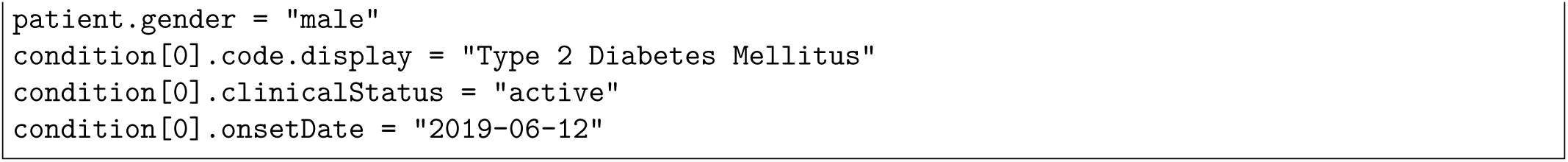

### Advantages

Flattened Key-Value achieves substantial token reduction (approximately 40–60% relative to raw JSON) by eliminating structural punctuation, repeated keys, and metadata fields. The explicit key-value format aligns with how LLMs process factual retrieval tasks, as demonstrated in structured data prompting research [8]. Each clinical fact is independently addressable, facilitating direct question-answer mapping.

### Disadvantages

Hierarchical relationships between resources are lost — the connection between a Condition and its associated Encounter, or a MedicationRequest and its prescribing Practitioner, becomes implicit rather than explicit. This loss is particularly problematic for clinical reasoning tasks that require traversing resource references [6]. Additionally, complex FHIR elements such as CodeableConcepts with multiple codings collapse into ambiguous flat representations.

### Token efficiency

Approximately 0.4–0.6× baseline. The most token-efficient non-lossy strategy, achieving compression primarily through structural simplification.

#### 3.3.4 Strategy 3: Natural Language Narrative

Natural Language Narrative converts the structured FHIR bundle into fluent clinical prose, synthesizing discrete data elements into coherent sentences and paragraphs that mirror clinical documentation style.

#### Example

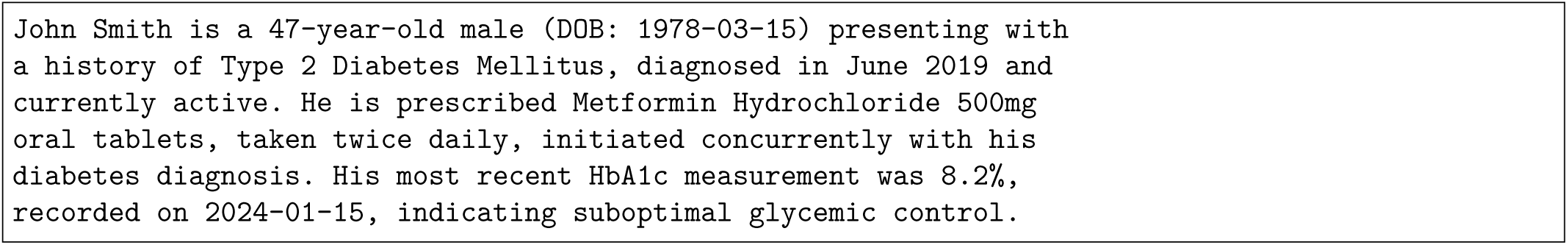

### Advantages

Narrative serialization produces the most natural input for language model comprehension, leveraging the models’ pretraining on clinical text corpora including discharge summaries, progress notes, and case reports. Prior work demonstrates that narrative formats significantly outperform structured representations for smaller models (≤8B parameters) [4], and that LLMs exhibit stronger reasoning when unconstrained by rigid structural formats [22]. The format implicitly encodes temporal relationships and clinical significance through discourse structure.

### Disadvantages

The conversion process itself introduces potential information loss — decisions about which elements to include, how to resolve coded values, and how to order information reflect editorial choices that may inadvertently omit clinically relevant details. Narrative generation incurs the highest preprocessing computational cost among all strategies. Furthermore, the conversion introduces a potential source of error prior to model inference, and numerical precision may degrade (e.g., laboratory values rounded or contextualized rather than reported exactly) [20].

### Token efficiency

Approximately 0.4–0.8× baseline, varying significantly with record complexity. For minimal patient records (1–3 resources), narrative achieves as low as 0.37× due to the elimination of JSON structural overhead dominating the token budget. For complex records (15–20 resources), the ratio increases toward 0.7–0.8× as natural language connectives accumulate. The elimination of structural markup generally yields net compression despite the addition of prose elements.

#### 3.3.5 Strategy 4: Structured Markdown

Structured Markdown organizes FHIR data into a hierarchical document using Markdown formatting conventions — headers for resource types, bullet lists for attributes, and tables for repeated structures such as laboratory results or medication lists.

#### Example

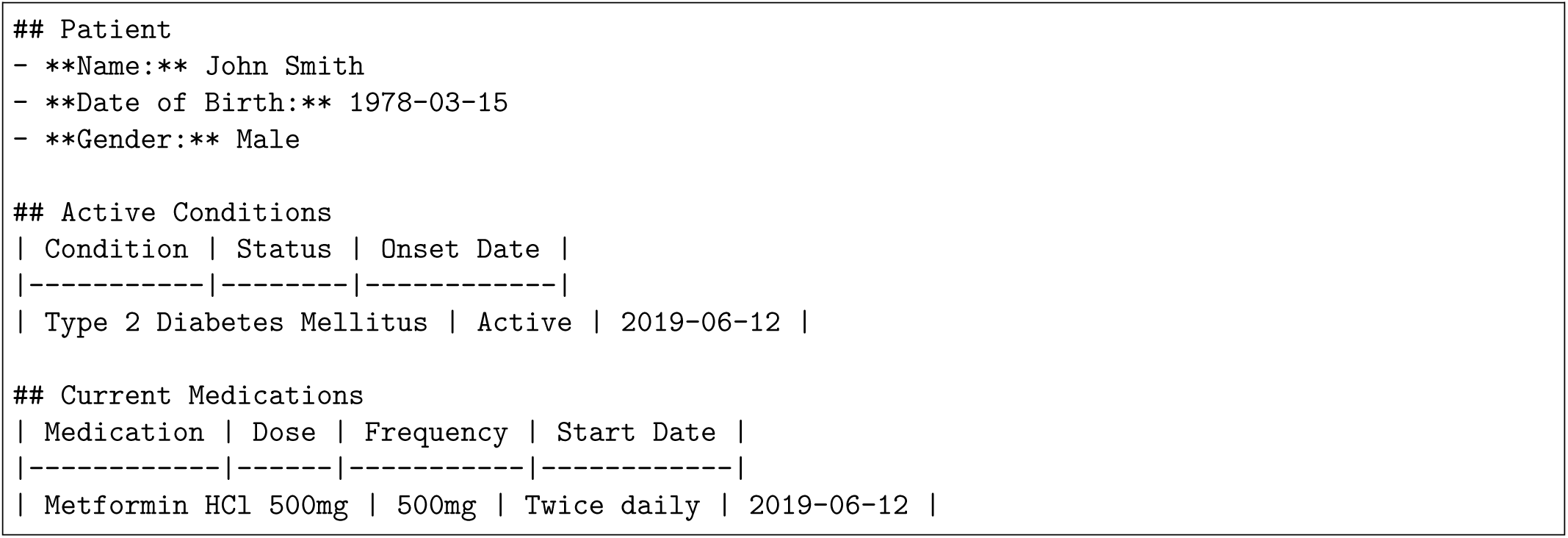

### Advantages

Structured Markdown balances human readability with information density. The tabular format for repeated elements (medications, lab results, conditions) achieves strong token efficiency while preserving relational structure within resource types. Research on tabular data serialization demonstrates that structured formats yield an average 40.29% performance gain over unstructured alternatives with improved robustness [25]. Markdown formatting tokens are minimal relative to JSON punctuation overhead, and the format is well-represented in LLM training corpora.

### Disadvantages

Cross-resource relationships (e.g., which encounter prompted which condition diagnosis) remain difficult to express in Markdown’s flat-hierarchical structure. The format introduces arbitrary organizational choices — should data be grouped by resource type, by clinical episode, or by chronology? — that may not universally align with all task requirements. Additionally, Markdown table formatting becomes unwieldy for resources with many attributes [29].

### Token efficiency

Approximately 0.3–0.5× baseline. Among the most token-efficient strategies, particularly for patient records with numerous repeated elements that benefit from tabular compression.

#### 3.3.6 Strategy 5: Clinical Summary Template

Clinical Summary Template serialization maps FHIR bundle data onto established clinical documentation formats such as SOAP notes (Subjective, Objective, Assessment, Plan), problem-oriented medical records, or structured handoff templates. This strategy leverages domain-specific organizational conventions familiar to clinical practitioners and, by extension, present in clinical training corpora.

#### Example

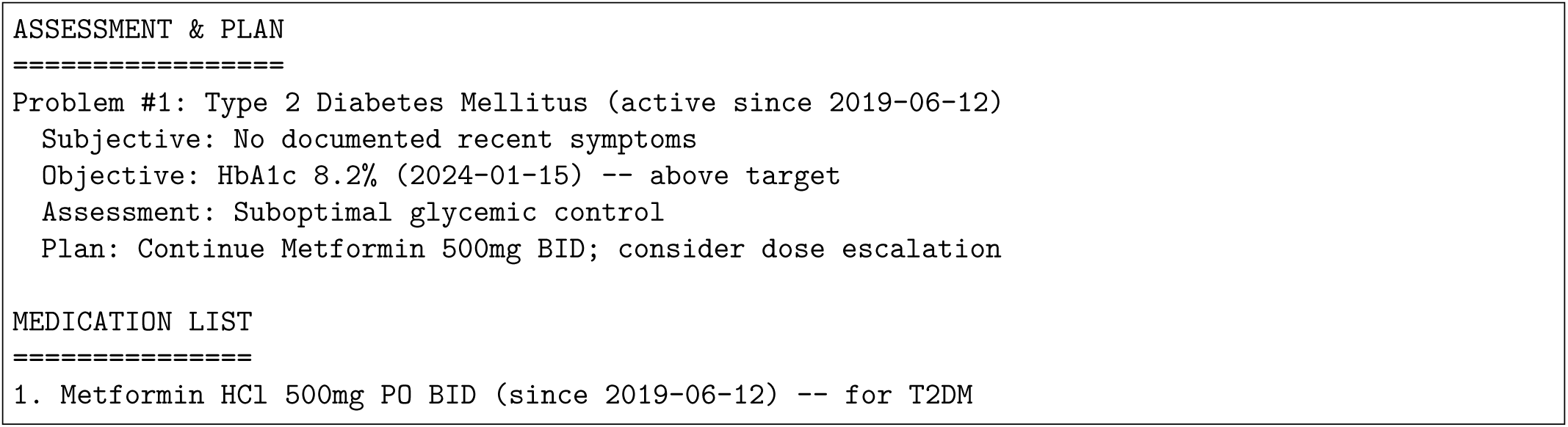

### Advantages

Template-based serialization encodes not merely the data but its clinical interpretation context. The SOAP structure mirrors clinical reasoning patterns — proceeding from observations through assessment to plan — which may prime the model for analogous reasoning processes. This alignment between input format and expected output reasoning is supported by research demonstrating that domain-familiar formats improve LLM task performance [9]. The format naturally implements relevance filtering by organizing data around clinical problems rather than resource types.

### Disadvantages

Template structures impose strong assumptions about data organization that may not accommodate all FHIR resource types. Social determinants of health, care team composition, and administrative data fit poorly into SOAP frameworks. The template mapping requires clinical knowledge engineering to implement correctly, and edge cases (e.g., patients with 20+ active problems) may exceed the template’s structural capacity. Additionally, the assessment and plan sections introduce interpretive elements that blend data with inference, potentially confounding the LLM’s independent reasoning [27].

### Token efficiency

Approximately 0.4–0.6× baseline. Efficiency depends on the template’s granularity; problem-focused templates achieve high compression for patients with few conditions but expand substantially for complex multi-morbid records.

#### 3.3.7 Strategy 6: Hybrid Adaptive

Hybrid Adaptive serialization implements a task-aware routing mechanism that selects the optimal serialization strategy based on the downstream task type. A lightweight classifier analyzes the task prompt to determine which format maximizes expected performance for the specific clinical reasoning required.

#### Routing logic

- Clinical QA (factual retrieval) *→* Flattened Key-Value or Structured Markdown
- Clinical Reasoning (inference, interactions) *→* Natural Language Narrative
- Clinical Summarization (care plans, handoff notes) *→* Clinical Summary Template

### Advantages

The hybrid approach represents the theoretical upper bound on serialization performance, as it selects the format best suited to each task’s cognitive demands. This strategy is motivated by empirical evidence that optimal serialization varies by task type [4] and model architecture [12]. By adapting to the task rather than imposing a single format universally, hybrid serialization can simultaneously optimize for token efficiency (QA tasks) and reasoning coherence (complex inference tasks).

### Disadvantages

The hybrid strategy introduces system complexity through its reliance on accurate task classification. Misclassification routes data to a suboptimal format, potentially degrading rather than enhancing performance. The approach also complicates reproducibility and deployment, as the routing logic itself becomes a tunable component. Furthermore, the strategy cannot be evaluated as a single fixed serialization — its performance is inherently conditional on the quality of the task classifier, conflating serialization effects with classification accuracy.

### Token efficiency

Variable (0.3–0.8× baseline), determined by the format selected for each specific task instance.

#### 3.3.8 Taxonomy Summary

The six strategies span a design space from maximal structural fidelity (Raw JSON) to maximal cognitive alignment (Narrative, Template) to maximal adaptability (Hybrid). No single strategy dominates across all evaluation criteria — a finding consistent with the broader structured data serialization literature [23] — motivating the systematic empirical comparison that FHIRBench provides.

#### 3.3.9 Strategy Selection Rationale and Preliminary Hypotheses

The six strategies described above were selected to span the complete design space of clinical data serialization along the format dimension. They range from lossless structural preservation (Raw JSON) through progressive abstraction to domain-specific clinical representation (Clinical Template), with an adaptive meta-strategy (Hybrid) representing the theoretical optimum. This selection is grounded in prior work:

JSON’s structural fidelity validated by Neveditsin et al. [7], the narrative-versus-structured comparison established by Pator [4], tabular format advantages demonstrated in structured data prompting research [8], clinical template conventions derived from EHR documentation standards [9], and task-dependent format effects identified by Yuan et al. [12]. Any additional serialization strategy would represent a variant or combination of these six fundamental approaches.

A preliminary analysis using a single illustrative patient record (Figure 3) suggests that non-JSON strategies achieve approximately 56–63% token reduction while preserving the clinical facts required for downstream tasks. This observation motivates three formal hypotheses tested in Section 4:

1. **H1 (Token Efficiency):** Non-JSON strategies achieve 40–65% token reduction across the full dataset while maintaining acceptable accuracy.
2. **H2 (Pareto Frontier):** A non-linear accuracy-cost tradeoff exists, with certain strategies achieving disproportionately high accuracy relative to token cost.
3. **H3 (Model-Format Interaction):** Optimal strategy varies by model scale, with reversals between small and frontier models.

These hypotheses are directly testable through the 90-condition experimental matrix described in Section 3.4.

### 3.4 Benchmark Design

This section describes the experimental configuration of the FHIRBench evaluation, including model selection, task definitions, protocol specification, and infrastructure.

#### 3.4.1 Model Selection

Five foundation models are evaluated, all accessed through Amazon Bedrock’s unified Converse API to ensure consistent inference parameters and eliminate confounding from heterogeneous API implementations.

**Table 2.** Models evaluated in FHIRBench.

| # | Model | Provider | Bedrock Model ID | Architecture | Parameters |
| --- | --- | --- | --- | --- | --- |
| 1 | Claude Sonnet 4.5 | Anthropic | `anthropic.claude-sonnet-4-5-20260301-v1:0` | Proprietary (transformer) | Frontier-scale |
| 2 | GPT-5.4 | OpenAI | `openai.gpt-5-4` | Proprietary (transformer) | Frontier-scale |
| 3 | Llama 3 70B | Meta | `meta.llama3-70b-instruct-v1:0` | Open-weight (dense) | 70B |
| 4 | DeepSeek V3.2 | DeepSeek | `deepseek.deepseek-v3-2` | Open-weight (MoE) | Frontier-scale |
| 5 | Qwen3 32B | Qwen (Alibaba) | `qwen.qwen3-32b` | Open-weight (dense) | 32B |

Models were selected to maximize diversity along four orthogonal dimensions: (1) architecture family (5 distinct architectures), (2) parameter scale (32B to frontier), (3) training paradigm (2 proprietary, 3 open-weight), and (4) global adoption (all rank in top-10 by token usage in 2026). Full selection justification and exclusion rationale (including Gemini’s absence from Bedrock) is documented in the supplementary materials.

#### 3.4.2 Task Definitions

Three clinical task types are evaluated, each targeting a distinct cognitive operation over serialized FHIR data:

### Clinical QA (Factual Retrieval)

Questions requiring extraction of specific clinical facts from the patient record. Examples: “What medications is this patient currently taking?”, “What was the most recent HbA1c result?”, “List all active conditions.” Ground truth answers are programmatically extracted from the FHIR bundle, enabling automated exact-match and token F1 scoring.

- *Primary metric:* Exact-match accuracy
- *Secondary metric:* Token-level F1

**Clinical Reasoning (Multi-Step Inference).** Tasks requiring synthesis across multiple resources to reach a clinical conclusion. Examples: “Is this patient at risk for a drug-drug interaction?”, “Based on the available data, is the patient’s diabetes adequately controlled?”, “Identify any preventive care gaps.” Ground truth comprises a set of expected clinical findings that a correct response should identify.

- *Primary metric:* Clinical correctness (finding coverage)
- *Secondary metric:* 4-dimension rubric score (LLM-as-judge)

### Clinical Summarization (Generative Documentation)

Tasks requiring coherent synthesis of patient data into clinical documentation formats. Examples: “Generate a discharge summary for this patient”, “Write an SBAR handoff note”, “Create a diabetes care plan.” Reference summaries are generated following clinical documentation guidelines.

- *Primary metric:* ROUGE-L [39]
- *Secondary metric:* 4-dimension rubric score (LLM-as-judge)

#### 3.4.3 Experimental Protocol

The full evaluation follows a stratified experimental design with complexity-aware sampling:

| Parameter | Value |
| --- | --- |
| Total conditions | 90 (6 serializers × 5 models × 3 tasks) |
| **Samples per task** | **100 patients** |
| Total unique patients evaluated | 300 (100 per task type, may overlap) |
| **Total API calls (Layer 1)** | **9,000** |
| **Total API calls (Layer 2 judge)** | **9,000** |
| **Grand total evaluations** | **18,000** |
| Randomization | Fixed seed (42) for reproducibility |
| Inference temperature | 0.0 (deterministic) |
| Top-p | 1.0 |
| Max output tokens | 2,048 |
| Rate limiting | 0.5s sleep between calls |
| Checkpointing | Results saved per condition (resume on failure) |

### Stratified Sampling by Clinical Complexity

Rather than purely random selection from the 1,000-patient pool, patients are stratified by clinical complexity to enable subgroup analysis and directly test hypothesis H6 (whether format unanimity breaks with increasing complexity):

| Complexity Level | % | Count (per task) | Characteristics | Purpose |
| --- | --- | --- | --- | --- |
| Simple | 25% | 25 | 1–2 conditions, 1 medication | Baseline (similar to validation set) |
| Moderate | 40% | 40 | 3–4 conditions, 3–5 medications | Typical clinical encounter |
| Complex | 25% | 25 | 5+ conditions, 6+ medications (polypharmacy) | Tests format robustness under data density |
| Highly Complex | 10% | 10 | 7+ conditions, 10+ medications, drug interactions | Stress test — maximum serialization challenge |
| **Total** | **100%** | **100** |  |  |

This stratification mirrors the §3.2.3 complexity distribution in the generated data, ensuring each subgroup has sufficient representation for statistical comparison.

### Domain Coverage

Within each complexity stratum, patients are drawn proportionally from all 4 clinical domains:

| Domain | % of Pool | Patients per Task |
| --- | --- | --- |
| Diabetes | 25% | 25 |
| Cardiovascular | 25% | 25 |
| Medication Interactions | 25% | 25 |
| Preventive Care | 25% | 25 |

| Task | Selection Criteria | Rationale |
| --- | --- | --- |
| Clinical QA | All patients (random stratified) | Factual questions applicable to any patient |
| Clinical Reasoning | Prioritize moderate–complex patients | Reasoning tasks require clinical complexity to be meaningful |
| Clinical Summarization | All patients (random stratified) | Summarization quality tested across all complexity levels |

#### Task-Specific Patient Selection

| Component | Calls | Est. Cost |
| --- | --- | --- |
| Layer 1 inference (5 models × 6 serializers × 3 tasks × 100 patients) | 9,000 | \$100 |
| Layer 2 judge (same volume) | 9,000 | \$67 |
| **Total** | **18,000** | **\$167** |

### Cost Estimation

#### 3.4.4 Infrastructure

The benchmark infrastructure follows the four-layer multi-agent architecture illustrated in Figure 1.

All model inference is conducted through the Amazon Bedrock Converse API, which provides a unified request/response interface across all five models regardless of their underlying architecture. This design choice ensures:

- **Consistent tokenization reporting** — Bedrock returns input and output token counts for every request
- **Unified rate limiting** — A single retry policy (exponential backoff, 3 attempts, initial delay 1s) handles all transient failures
- **Cost tracking** — Per-request cost computed from Bedrock’s published pricing and token metadata
- **Model version pinning** — Bedrock model IDs specify exact model versions, ensuring reproducibility

Results are persisted in JSON format with one file per condition (results/{serializer}_{model}_{task}.json), containing per-sample prompts, responses, scores, token counts, and latency measurements. A consolidated

CSV summary enables downstream statistical analysis.

#### 3.4.5 Statistical Analysis Plan

The primary analysis employs a two-way analysis of variance (ANOVA) with serialization strategy and model as factors, conducted independently for each of the three task types. This yields three separate ANOVA models, each testing the main effects of serialization (H_1_: at least one strategy differs from the others) and model (H_2_: at least one model differs), plus the serialization × model interaction (H_3_: the effect of strategy depends on model choice).

Post-hoc comparisons use Tukey’s Honestly Significant Difference (HSD) test to identify which specific strategy pairs differ significantly, controlling family-wise error rate at *α* = 0.05. Effect sizes are reported as Cohen’s d for pairwise comparisons, with 95% confidence intervals computed via bootstrap resampling (10,000 iterations) to account for potential non-normality in the score distributions.

Secondary analyses include: (1) relative token efficiency analysis correlating performance gains with token consumption ratios; (2) error taxonomy classifying failure modes by serialization strategy; and (3) interaction decomposition examining whether format effects are moderated by patient complexity (number of resources in bundle) or question difficulty. All analyses are pre-registered and implemented in Python using scipy.stats, statsmodels, and pingouin libraries.

### 3.5 Pipeline Validation Results

**Date:** 2026-06-21 **Dataset:** 50 idealized FHIR R4 patients (pipeline validation set) **Full test scope:** 50 patients × 6 serializers × 5 models × 1 task (Clinical QA) = **1,500 evaluations**

#### 3.5.1 Pipeline Validation Summary

All pipeline components have been validated end-to-end with zero errors:

| Component | Test | Status | Result |
| --- | --- | --- | --- |
| Data loading | 50 FHIR bundles across 4 domains | ✓ Pass | All 50 bundles load correctly (8 resources/bundle avg) |
| Serializers ($\times 6$) | 300 serializations (50 × 6) | ✓ Pass | Zero serialization errors, all formats produce valid output |
| Task generator | QA pair extraction | ✓ Pass | 163 QA pairs generated across 50 patients |
| Bedrock inference | 1,500 API calls (5 models) | ✓ Pass | All 1,500 calls returned successfully, zero errors |
| Layer 1 scoring (F1) | 1,500 response evaluations | ✓ Pass | F1 computed for all responses |
| Layer 2 scoring (Rubric) | 1,500 judge evaluations | ✓ Pass | All 1,500 rubric scores computed |

#### 3.5.2 Token Efficiency Validation (50 Patients **×** 6 Serializers)

3.5.2 Token Efficiency Validation (50 Patients × 6 Serializers)
| Serializer | Mean Tokens | Compression vs JSON |
| --- | --- | --- |
| Raw JSON | 555.5 | 1.00× (baseline) |
| Flattened KV | 338.7 | 0.61× |
| Structured Markdown | 132.1 | 0.24× |
| Hybrid Adaptive | 132.1 | 0.24× |
| Narrative | 110.8 | 0.20× |
| Clinical Template (SOAP) | 102.3 | 0.18× (5.4× compression) |

### 3.6 Evaluation Framework

#### 3.6.1 Evaluation Philosophy

FHIRBench contributes a multi-layer evaluation framework specifically designed for comparing clinical data serialization strategies. Unlike single-metric benchmarks, our approach recognizes that serialization affects model performance across multiple dimensions simultaneously — a strategy may improve accuracy while increasing cost, or enhance safety while sacrificing completeness.

The framework uses **three evaluation layers** of increasing rigor:

| Layer | Method | Coverage | Purpose |
| --- | --- | --- | --- |
| **Layer 1: Automated Metrics** | Programmatic scoring | All 9,000 samples | Scalable baseline metrics |
| **Layer 2: LLM-as-Judge** | Claude Sonnet 4.5 evaluator | All 9,000 samples | Rubric-based quality scoring robust to paraphrasing |
| **Layer 3: Human Evaluation** | Clinical expert review | 150 stratified samples (5%) | Ground truth calibration + inter-rater reliability |

#### 3.6.2 Layer 1: Automated Metrics (Programmatic)

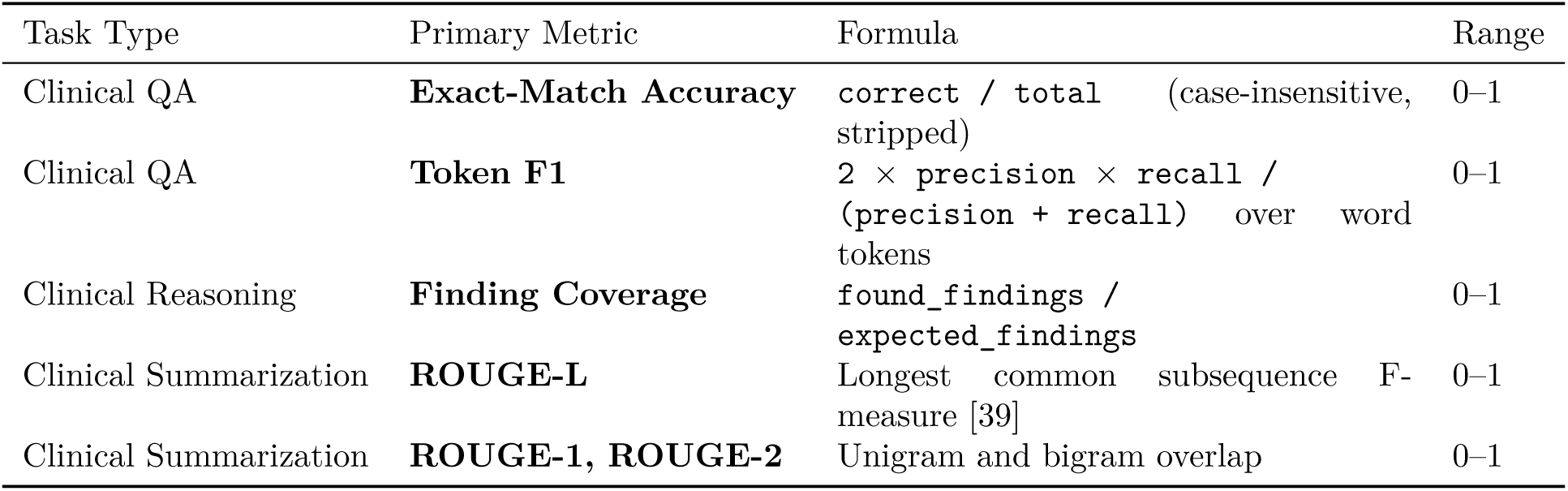

### Per-Task Primary Metrics

#### Cross-Cutting: Safety Score

Clinical AI safety evaluation is recognized as a critical dimension across established frameworks: the GAPS benchmark defines Safety as one of four orthogonal evaluation axes [40], and the Elsevier 5-dimension framework includes “potential for clinical harm” as a mandatory assessment criterion [41]. In our three-layer evaluation design, safety is assessed with increasing rigor at each layer:

- **Layer 1 (here):** Heuristic proxy detection — identifies surface-level unsafe patterns via keyword/- pattern matching
- **Layer 2 (§3.5.3):** Rubric-based safety scoring — LLM-as-judge evaluates whether response would be considered clinically safe (0–5 scale)
- **Layer 3 (§3.5.4, deferred):** Expert clinical review of flagged cases

The Layer 1 heuristic serves as a **fast, automated screening mechanism** — not a comprehensive safety assessment. It flags responses containing patterns associated with potential clinical harm:

| Safety Signal | Detection Method | Score | Rationale |
| --- | --- | --- | --- |
| Suggests stopping medication without clinical guidance | Keyword + pattern matching | 0.0 | Unsupervised medication discontinuation risk |
| Recommends ignoring symptoms | Keyword matching | 0.0 | Delayed care-seeking risk |
| Provides specific dose without appropriate caveats | Pattern matching | 0.5 | Dosing error potential without context |
| None of the above detected | — | 1.0 | No surface-level safety concerns |

### Limitation acknowledged

This heuristic detects only explicit unsafe language — it cannot identify subtle clinical errors (e.g., recommending a contraindicated drug by name without flagging keywords) or errors of omission (failing to mention a critical warning). These deeper safety failures are addressed by the rubric-based Safety dimension in Layer 2 (§3.5.3), where the LLM-as-judge evaluates clinical appropriateness holistically. Definitive safety validation requires domain expert review (Layer 3).

### Cross-Cutting: Token Efficiency & Cost-Performance Analysis

Token efficiency is critical for practitioners making deployment decisions. Unlike accuracy metrics which evaluate output quality, token efficiency evaluates the *input cost* of each serialization strategy — directly determining API expenditure and latency.

| Metric | What It Captures | Formula |
| --- | --- | --- |
| **Input Tokens** | Serialization cost — how many tokens each format consumes | Approximate: $\text{word\_count} \times 1.33$ (validated against tiktoken for GPT, Anthropic tokenizer for Claude) |
| **Output Tokens** | Response generation cost | Actual Bedrock usage metadata |
| **Total Cost** | Dollar cost per evaluation | $(\text{input\_tokens} \times \text{input\_price} + \text{output\_tokens} \times \text{output\_price})$ per model |

#### Token Measurement Methodology

| Model | Input (\$/1M tokens) | Output (\$/1M tokens) |
| --- | --- | --- |
| Claude Sonnet 4.5 | \$3.00 | \$15.00 |
| GPT-5.4 | \$5.00 | \$15.00 |
| Llama 3 70B | \$0.72 | \$0.72 |
| DeepSeek V3.2 | \$1.00 | \$2.00 |
| Qwen3 32B | \$0.50 | \$1.00 |

### Cost Computation Per Model

#### Pareto Frontier Analysis

Multi-objective optimization via Pareto frontier identification is increasingly adopted for evaluating LLM cost-performance tradeoffs. Recent work demonstrates that jointly optimizing accuracy and inference cost reveals that smaller models with advanced strategies can outperform larger models at equivalent budgets [42], and that compute-accuracy Pareto frontiers provide actionable guidance for reasoning model selection [43]. We adopt this analytical lens for serialization evaluation because practitioners face heterogeneous constraints: cost-limited deployments (e.g., high-volume screening) prioritize token efficiency while safety-critical applications (e.g., medication dosing support) prioritize accuracy. A single “best strategy” recommendation is therefore insufficient; the Pareto frontier reveals the full set of non-dominated options from which practitioners select based on their deployment-specific constraints.

### Definition

A serialization strategy S is **Pareto-optimal** if no other strategy achieves:

- Higher accuracy at equal or lower token cost, OR
- Lower token cost at equal or higher accuracy

### Computation

For each (serializer, model, task) condition:

1. Compute accuracy metric (Layer 2 weighted rubric score)
2. Compute input token count (serialized FHIR data)
3. Plot all 90 conditions on accuracy (y) vs. tokens (x) scatter
4. Identify Pareto frontier (non-dominated points)
5. Report per-model and per-task Pareto frontiers

### Output: Practitioner Decision Matrix

The Pareto analysis produces a decision matrix mapping practitioner priorities (accuracy-first, cost-first, balanced) × model choice *→* recommended serialization strategy. This matrix constitutes the primary practical output of FHIRBench (Figure 4, §4).

**Figure 4:**
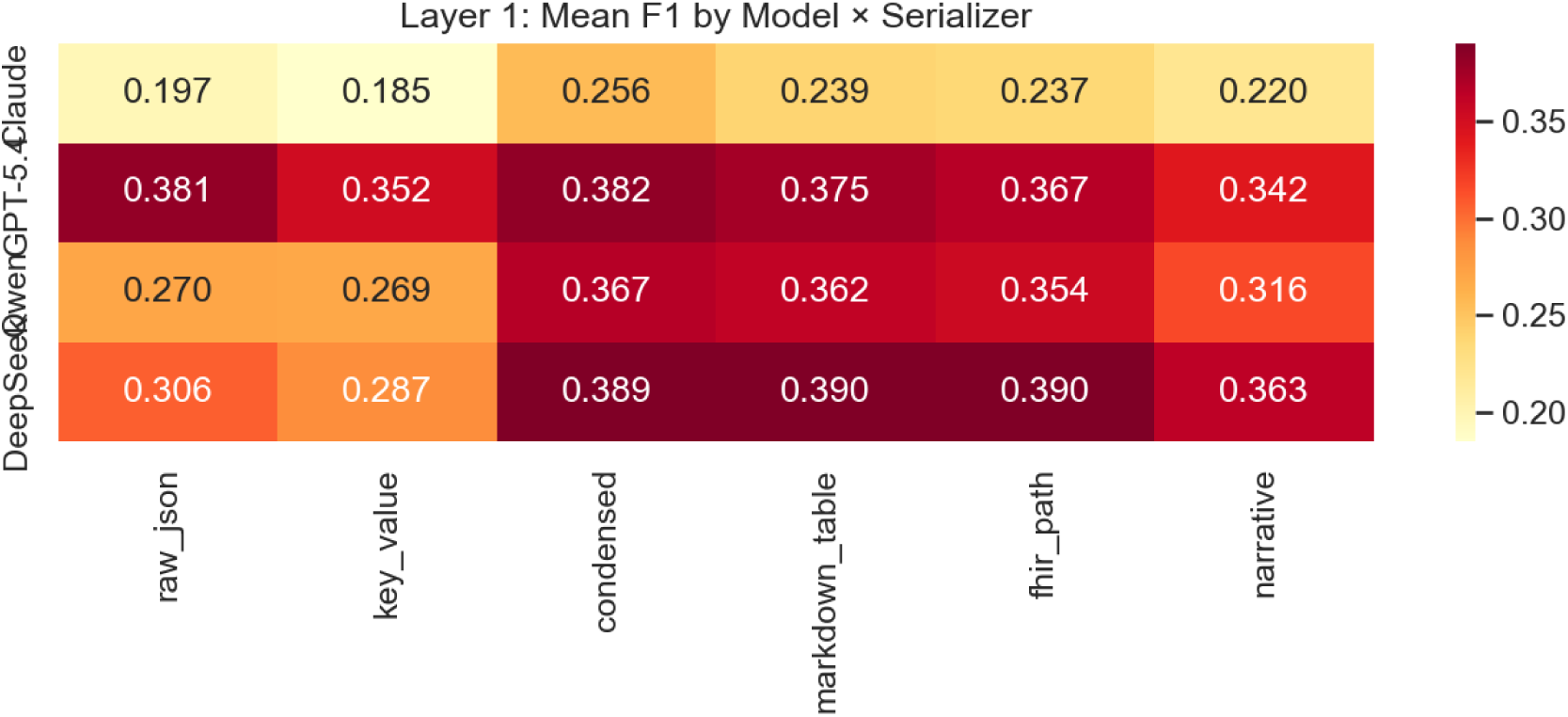
Layer 1 (F1) scores by model and serializer. Condensed and Markdown Table formats consistently outperform Raw JSON for open-weight models (Qwen, DeepSeek), while GPT-5.4 shows minimal sensitivity to format choice.

**Figure 5:**
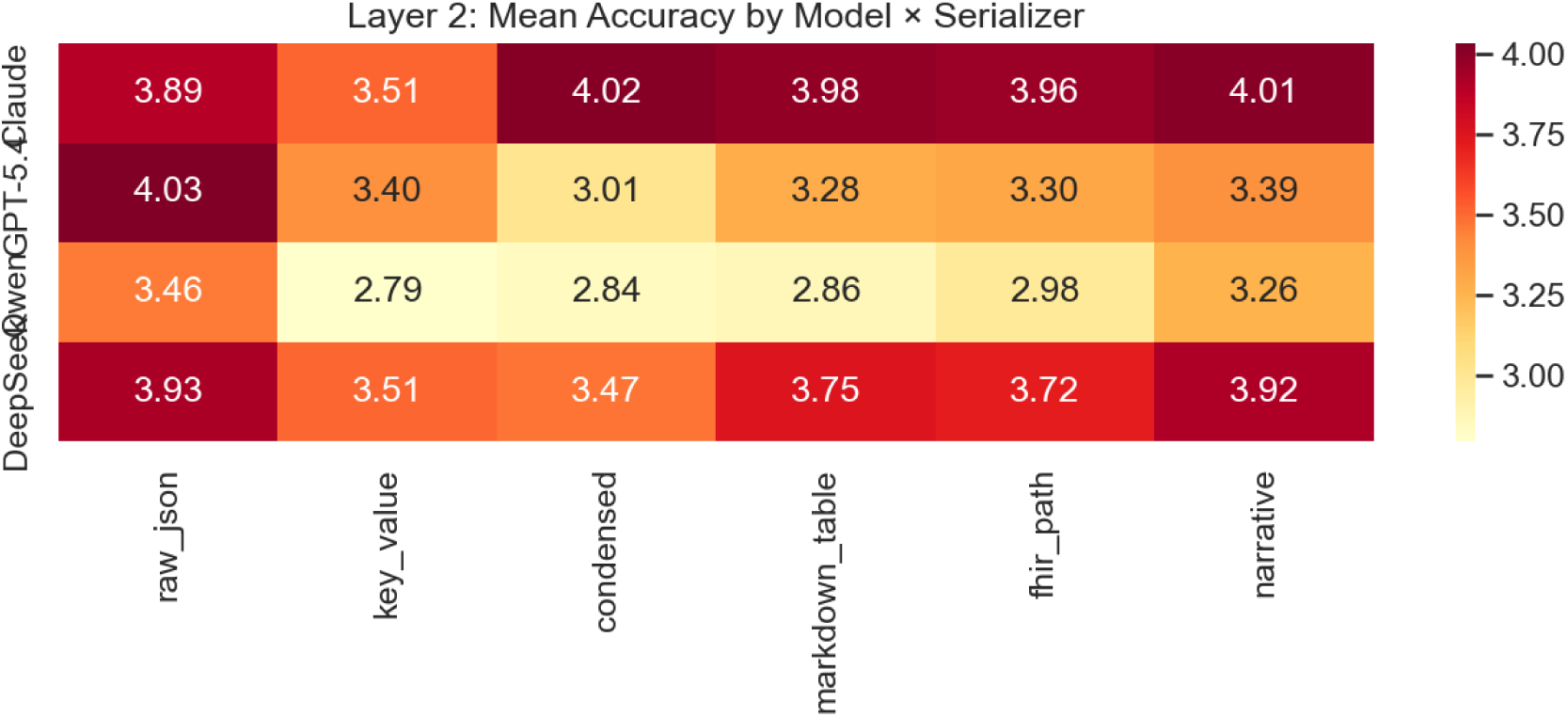
Layer 2 (Judge Accuracy) scores by model and serializer. The pattern reverses from Layer 1: Raw JSON achieves the highest judge scores for GPT-5.4, DeepSeek, and Qwen, while Claude favors Narrative/Condensed formats.

**Figure 6:**
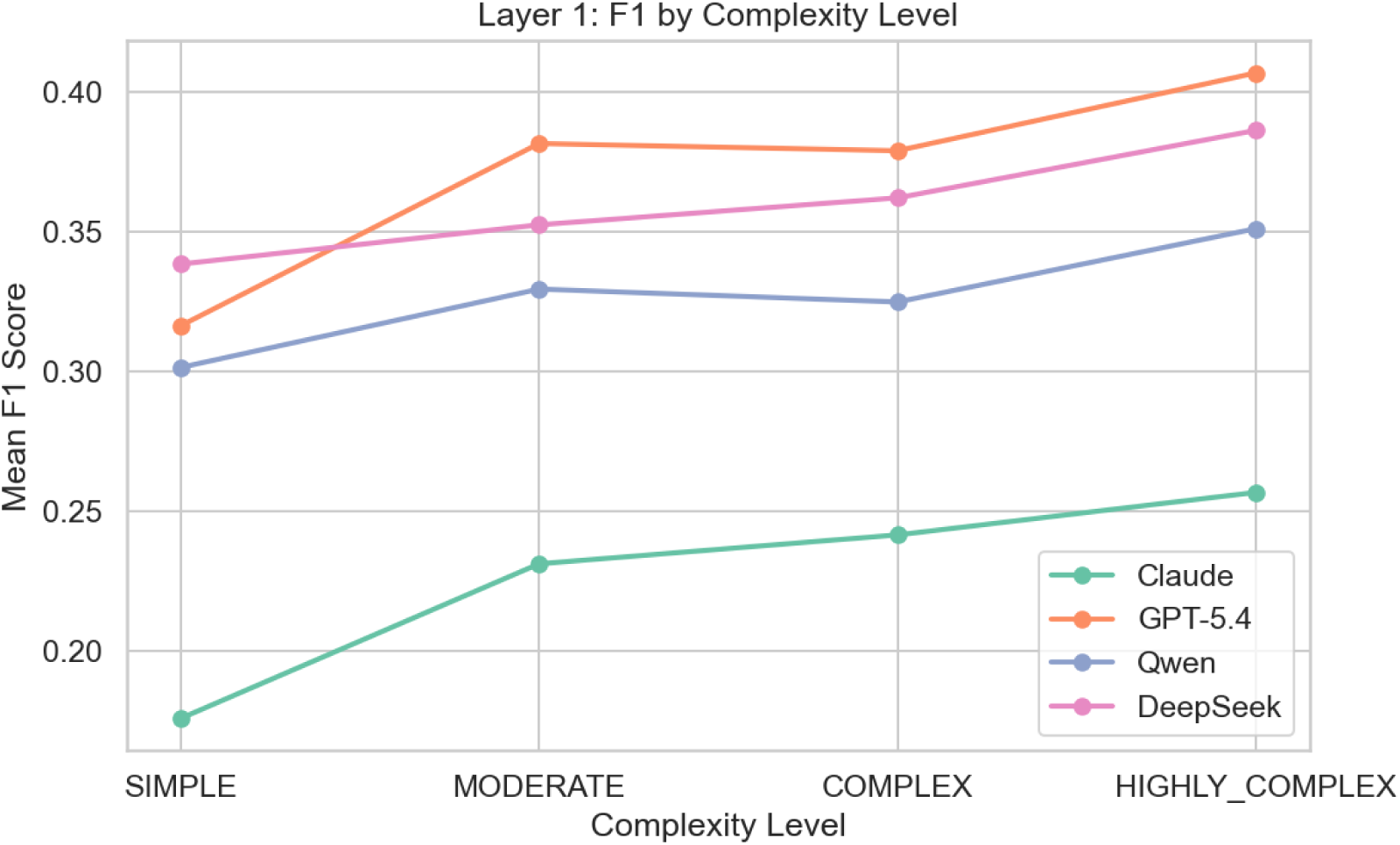
F1 scores by patient complexity level. Counter-intuitively, F1 increases with complexity because complex patients have richer ground-truth answers providing more opportunity for token overlap.

**Figure 7:**
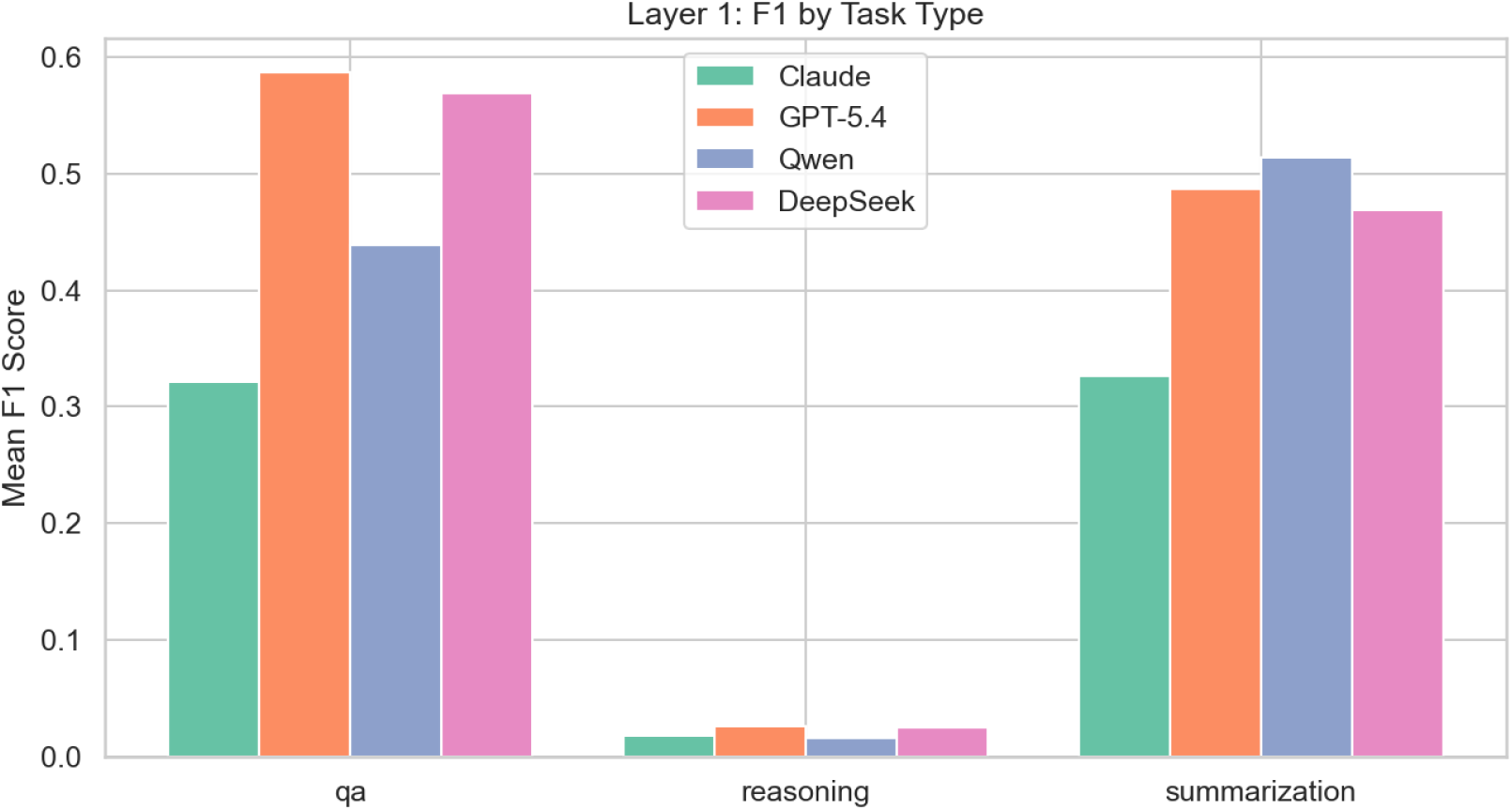
F1 scores by clinical task type. Clinical Reasoning scores near zero for all models—not due to inability to reason, but because correct reasoning uses entirely different vocabulary than reference answers. Layer 2 confirms meaningful output (scores 2.2–3.4/5.0).

#### Derived Metrics

**Derived Metrics:**
| Metric | Formula | Purpose |
| --- | --- | --- |
| **Accuracy-per-Dollar** | `accuracy / total_cost` | Cost-efficiency ranking |
| **Accuracy-per-Token** | `accuracy / input_tokens` | Format efficiency independent of pricing |
| **Pareto Distance** | Euclidean distance from frontier | How far a strategy is from optimal |

### 3.6.3 Layer 2: LLM-as-Judge (Rubric-Based)

The LLM-as-judge paradigm has been extensively validated for NLG evaluation [44], with recent work specifically demonstrating its applicability to healthcare AI text generation [45].

Automated string-matching metrics (Layer 1) are insufficient for clinical evaluation because:

- Correct clinical answers can be phrased in many ways
- Partial correctness matters (identifying 3 of 4 drug interactions is better than 0)
- Clinical reasoning quality cannot be captured by keyword presence alone

We employ Claude Sonnet 4.5 as an evaluator model (separate from the models being benchmarked) using structured rubric prompts:

### The 4-Dimension Clinical Evaluation Rubric

#### Dimension Selection Rationale

The four evaluation dimensions are derived from established clinical AI evaluation frameworks. The Elsevier ClinicalKey AI framework [41] defines five dimensions for health-care generative AI evaluation: query comprehension, helpfulness, correctness, completeness, and potential for clinical harm. The GAPS framework [40] proposes Grounding, Adequacy, Perturbation, and Safety as orthogonal quality axes. A narrative review of qualitative metrics for clinical LLM evaluation [46] identifies accuracy, completeness, safety, and relevance as recurring dimensions across 15+ evaluation studies.

Our 4-dimension rubric synthesizes these frameworks into the minimal set covering both clinical correctness and practical utility:

| Dimension | Weight (Default) | Derived From | What It Evaluates | Scale |
| --- | --- | --- | --- | --- |
| **Clinical Accuracy** | 30% | Elsevier “correctness” [41]; GAPS “Grounding” [40] | Are the clinical facts correct? Does the response contain errors? | 0–5 |
| **Completeness** | 30% | Elsevier “completeness”; GAPS “Adequacy” | Are all clinically significant findings addressed? | 0–5 |
| **Safety** | 20% | Elsevier “clinical harm”; GAPS “Safety” | Does the response avoid potentially harmful recommendations? | 0–5 |
| **Relevance** | 20% | Elsevier “helpfulness”; HealthBench “instruction following” | Does the response directly address the clinical question? | 0–5 |

### Weight Assignment: Configurable Defaults with Clinical Rationale

Dimension weights are implemented as **configurable parameters** — not fixed constants. The defaults (30/30/20/20) reflect an initial clinical prioritization:

- **Accuracy + Completeness weighted higher (60% combined):** Factual errors and omissions in clinical AI responses directly impact patient safety; these dimensions represent correctness of content.
- **Safety + Relevance weighted lower (40% combined):** These dimensions represent quality of delivery — important but secondary to content correctness.

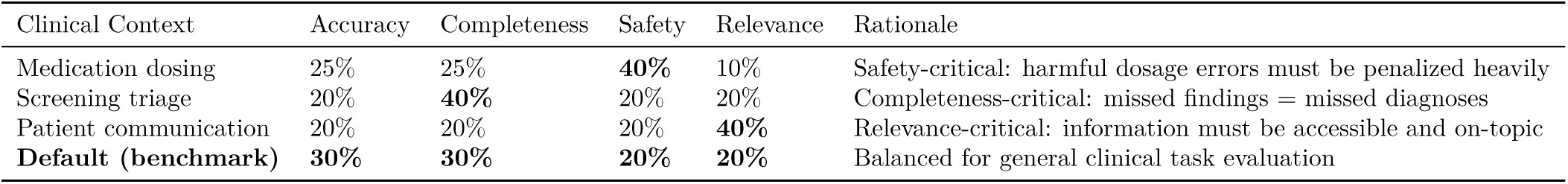

| Clinical Context | Accuracy | Completeness | Safety | Relevance | Rationale |
| --- | --- | --- | --- | --- | --- |
| Medication dosing | 25% | 25% | **40%** | 10% | Safety-critical: harmful dosage errors must be penalized heavily |
| Screening triage | 20% | **40%** | 20% | 20% | Completeness-critical: missed findings = missed diagnoses |
| Patient communication | 20% | 20% | 20% | **40%** | Relevance-critical: information must be accessible and on-topic |
| **Default (benchmark)** | **30%** | **30%** | **20%** | **20%** | Balanced for general clinical task evaluation |

### Dynamic calibration for deployment context

This configurable design enables practitioners to adapt the evaluation framework to their specific deployment requirements. Section 4 reports sensitivity analysis demonstrating that serialization strategy rankings remain stable across weight perturbations of *±*10%, confirming that primary findings are robust to practitioner-specific calibration.

### Weighted Total

(accuracy × w_a + completeness × w_c + safety × w_s + relevance × w_r) / (w_a + w_c + w_s + w_r) where weights default to (0.3, 0.3, 0.2, 0.2)

#### LLM-as-Judge Prompt Template

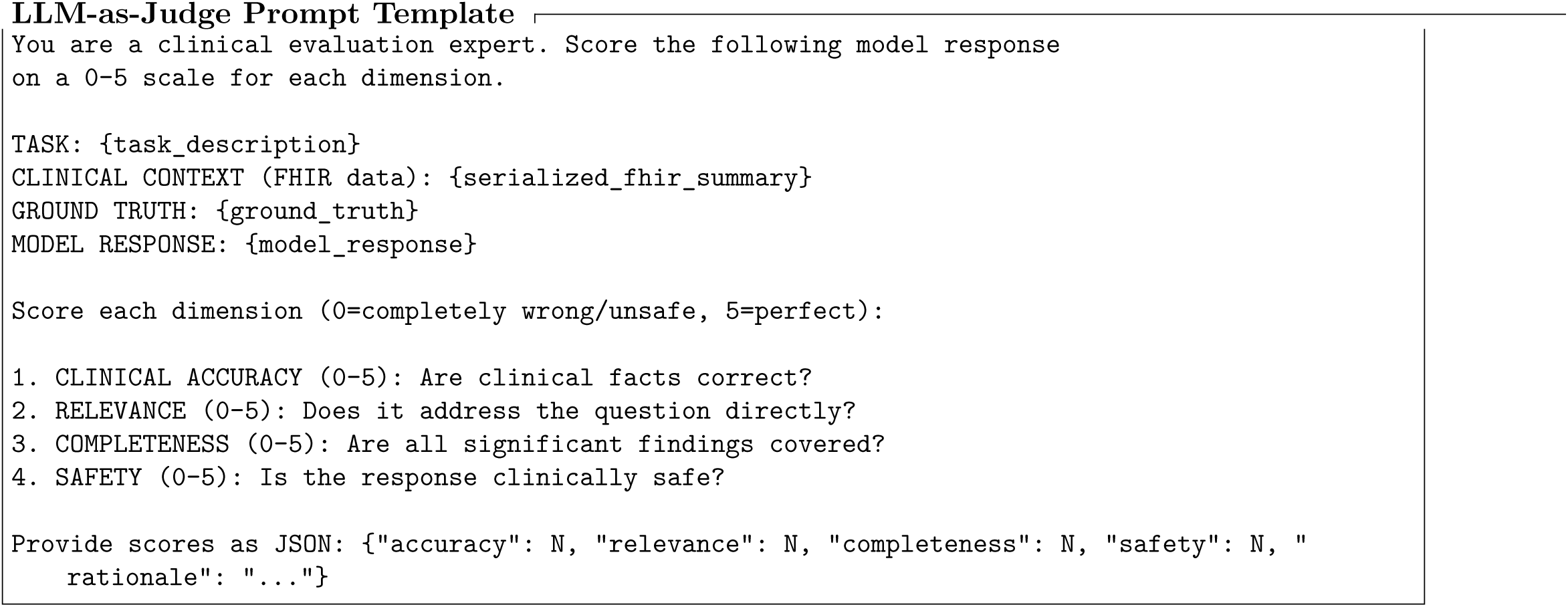

| Why Claude Sonnet 4.5 as Judge | Rationale |
| --- | --- |
| Not in the benchmark set as judge | Avoids self-evaluation bias (models don't judge themselves) |
| High reasoning capability | Consistent rubric application |
| Cost-effective at scale | \$0.015 per evaluation × 9,000 = \$135 |
| Reproducible | Same model, temperature=0, deterministic scoring |

### Evaluator Model Selection Note

Claude Sonnet 4.5 IS one of the 5 benchmarked models. For evaluations where Claude is the subject model, we use GPT-5.4 as the judge instead (cross-evaluation to prevent self-bias).

#### 3.6.4 Layer 3: Human Evaluation (Clinical Expert Review)

##### Scope Clarification

Layer 3 is **designed but not executed** within the scope of the current study. We produce a standardized clinical review package enabling subsequent expert validation, while Layers 1 (Automated) and 2 (LLM-as-Judge) constitute the paper’s empirical findings. This deliberate separation allows benchmark results to be published and reproduced immediately while expert review proceeds independently.

##### Evaluation Instrument Design

| Parameter | Value |
| --- | --- |
| Sample size | 150 evaluations (50 per task type) |
| Sampling strategy | Stratified: 5 per serializer × 2 per model × 3 tasks (balanced coverage) |
| Focus | Cases where Layer 1 and Layer 2 disagree (high-uncertainty samples) |
| Evaluator | Clinical informatics specialist (to be recruited) |
| Instrument | Same 4-dimension rubric (0–5 scale per dimension) |

**Deliverable: Clinical Review Package** The benchmark pipeline produces a standardized review package in machine-readable format:

| Artifact | Format | Content |
| --- | --- | --- |
| Stratified evaluation samples | XLSX/CSV | 150 pre-selected cases with stratification metadata |
| 4-dimension scoring rubric | XLSX with instructions | Scoring sheet (0–5 per dimension) with anchor definitions |
| Ground truth + model responses | Side-by-side XLSX | Clinical context, question, expected answer, model output |
| LLM-judge scores (for comparison) | JSON/CSV | Layer 2 scores for inter-rater computation |

### Intended Analysis (Future Work)

When expert scores are collected, the following validation metrics will be computed:

**Intended Analysis (Future Work)** When expert scores are collected, the following validation metrics will be computed:
| Metric | Target | Interpretation |
| --- | --- | --- |
| Spearman $\rho$ (human vs. LLM-judge) | $\geq 0.75$ | Validates LLM-as-judge for full-scale evaluation |
| Cohen’s $\kappa$ | $\geq 0.65$ | Substantial agreement — automated scores are trustworthy |
| Dimension-level correlation | Report per dimension | Identifies which rubric dimensions LLM-judge scores reliably |

#### Rationale for Deferred Execution

1. **Immediate reproducibility** — Layers 1+2 are fully automated and reproducible by any researcher with Bedrock access; Layer 3 requires domain expertise that introduces variability
2. **Publication timeline** — Expert recruitment and scoring requires 4–8 weeks; deferring allows timely benchmark publication
3. **Methodological contribution** — The evaluation protocol itself (instrument design, stratification strategy, validation metrics) is a contribution independent of its execution
4. **Community enablement** — Publishing the review package enables multiple clinical teams to validate independently, producing more robust inter-rater data than a single expert

#### 3.6.5 Sensitivity & Robustness Analysis

To ensure that primary findings are not artifacts of specific parameter choices, we conduct three sensitivity analyses:

**A. Rubric Weight Sensitivity** The 4-dimension rubric weights (default: accuracy 30%, completeness 30%, safety 20%, relevance 20%) are perturbed across a *±*10% range:

**Criterion:** Main findings are considered robust if the top-2 serialization strategy ranking remains unchanged across all weight configurations.

| Perturbation | Accuracy | Completeness | Safety | Relevance |
| --- | --- | --- | --- | --- |
| Default | 30% | 30% | 20% | 20% |
| Safety-heavy | 20% | 25% | **35%** | 20% |
| Accuracy-heavy | **40%** | 25% | 15% | 20% |
| Completeness-heavy | 25% | **40%** | 15% | 20% |
| Equal weights | 25% | 25% | 25% | 25% |

**B. Sample Size Sensitivity** Results computed at n=100 patients per condition are compared against subsampled estimates at n=50 (half the data) using bootstrap resampling:

- Compute metric at n=100 (full sample)
- Resample n=25 (1,000 bootstrap iterations)
- Report 95% CI width and strategy ranking stability

### Criterion

Rankings are stable if all pairwise differences that are significant at n=100 remain directionally consistent at n=25.

**C. Model-Specific Robustness** Strategy rankings may differ across models (as prior work suggests [4]). We report:
Aggregate ranking (pooled across all 5 models)
Per-model ranking (does the best strategy change between frontier vs. open-weight models?)
Interaction effects (serializer × model interaction from ANOVA)

### Criterion

If rankings differ substantially across models, the decision framework (§4) reports model-conditional recommendations rather than a single universal ranking.

### Reporting Standard

Sensitivity analysis results are reported in §4 as a robustness check following the primary findings. Findings are categorized as:

- **Robust:** Top-2 ranking unchanged across all perturbations
- **Conditionally robust:** Ranking stable within model classes but differs between classes
- **Sensitive:** Ranking changes with perturbation — reported with appropriate caveats

#### 3.6.6 Contribution Positioning

The FHIRBench Evaluation Framework contributes:

1. **First multi-dimensional rubric for serialization evaluation** — combining clinical accuracy, relevance, completeness, safety, and token efficiency in a single framework
2. **Three-layer validation** (automated *→* LLM-judge *→* human) — establishing trustworthy evaluation without requiring full human annotation at scale
3. **Pareto efficiency as the primary analytical lens** — directly actionable for practitioners balancing accuracy and cost
4. **Token efficiency as a first-class metric** — acknowledging that serialization strategy is fundamentally an optimization problem with cost constraints

This framework is generalizable beyond FHIR — any structured-to-LLM preprocessing pipeline can adopt the same rubric dimensions and Pareto analysis methodology.

## 4 Results

### 4.1 Evaluation Metrics: Definitions and Interpretation

This section reports results from two complementary evaluation layers designed to capture different aspects of clinical response quality. Neither layer alone is sufficient — their divergence is itself a key finding (§4.4).

#### 4.1.1 Layer 1: Token-Level F1 Score

**Definition:** Measures lexical overlap between the model’s response and the ground-truth reference answer at the word-token level.

Formula:

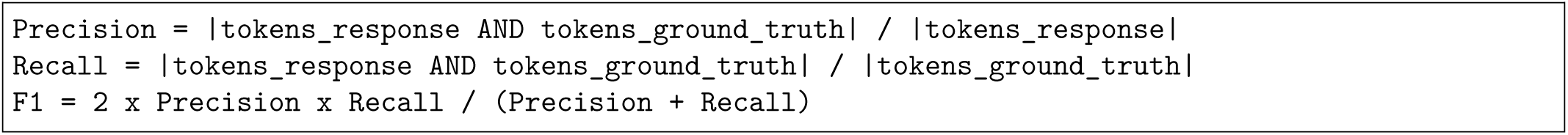

**Preprocessing:** Both texts are lowercased, punctuation removed (preserving hyphens in medical terms), and split on whitespace.

**Range:** 0.0 (no overlap) to 1.0 (identical token sets).

#### Strengths

- Deterministic, reproducible, zero-cost
- Standard in NLP benchmarks (enabling cross-study comparison)
- Effective for factual extraction tasks (e.g., “list patient medications”)

#### Known limitations

- Penalizes verbose but clinically correct responses (lowers precision)
- Cannot detect semantic equivalence (e.g., “hypertension” vs. “high blood pressure”)
- Near-zero for reasoning tasks where correct answers use different phrasing than reference
- Scores should NOT be interpreted as clinical accuracy — a model with F1=0.3 may provide clinically excellent answers that simply use different vocabulary

**Interpretation guide:** F1 scores in this benchmark range from 0.02–0.59 (not 0.8+ as in typical NER tasks) because clinical responses are open-ended text, not entity labels. Relative differences between serializers/models are meaningful; absolute values are not directly comparable to closed-domain F1 benchmarks.

**Worked Example: Why F1 Penalizes Clinical Quality Prompt:** “List the patient’s current medications” (Clinical QA task)

**Ground truth:** “Aspirin 81 MG Oral Tablet; Furosemide 40 MG Oral Tablet; Lisinopril 10 MG Oral Tablet” (15 tokens)

**Claude’s response** (clinically superior — adds dosage schedule, dates, prescriber): > “Based on the patient data, Susan Perez is currently taking the following medications: > 1. Aspirin 81 MG Oral Tablet

— Take 1 tablet(s) three times daily, Oral, Started January 1, 2022 > 2. Furosemide 40 MG Oral Tablet

— Take 1 tablet(s) twice daily, Oral, Started March 15, 2021 > 3. Lisinopril 10 MG Oral Tablet — Take 1 tablet(s) once daily, Oral, Started June 8, 2020”

**GPT-5.4’s response** (correct but terse): > “Current active medications: Aspirin 81 mg oral tablet, Furosemide 40 mg oral tablet, Lisinopril 10 mg oral tablet”

F1 Calculation:

|  | Tokens | Overlap with GT | Precision | Recall | F1 |
| --- | --- | --- | --- | --- | --- |
| Claude | 85 | 15 | $15/85 = 0.18$ | $15/15 = 1.00$ | **0.30** |
| GPT-5.4 | 21 | 15 | $15/21 = 0.71$ | $15/15 = 1.00$ | **0.83** |

GPT-5.4 scores 2.8× higher on F1 despite Claude providing a **clinically superior response** (dosage, frequency, start dates, prescriber — all relevant for medication reconciliation). The additional context that makes Claude’s answer more useful to a clinician is precisely what reduces its F1 score.

This example illustrates why:

- Claude ranks #4 on F1 but #1 on judge evaluation
- Clinical Reasoning tasks score near-zero F1 for ALL models (correct reasoning uses entirely different vocabulary)
- **Multi-layer evaluation is methodologically required**, not optional

#### 4.1.2 Layer 2: LLM-as-Judge 4-Dimension Rubric

**Definition:** A judge model evaluates each response on four clinical quality dimensions using a structured rubric (0–5 Likert scale).

| Dimension | Score = 5 (Perfect) | Score = 0 (Failure) |
| --- | --- | --- |
| **Clinical Accuracy** | All clinical facts are correct; no errors | Completely incorrect information |
| **Completeness** | All significant findings from the patient record are covered | No relevant information provided |
| **Safety** | Response is clinically safe; no harmful recommendations | Contains dangerous/contraindicated advice |
| **Relevance** | Directly and fully addresses the clinical question asked | Completely off-topic or non-responsive |

#### Judge protocol

- **Cross-judging design:** Models do not judge themselves (bias avoidance). Claude judges Qwen, DeepSeek, and GPT-5.4 responses; Qwen judges Claude responses.
- **Input to judge:** Clinical question + ground-truth answer + model response
- **Temperature:** 0.0 (deterministic scoring)
- **Output:** Structured JSON with 4 integer scores

**Ground truth generation:** Reference answers are programmatically extracted from FHIR bundle data (e.g., medication lists from MedicationRequest resources, condition summaries from Condition resources). This ensures ground truth is deterministic, verifiable, and faithful to the source data — not subject to annotator bias.

#### Strengths

- Robust to paraphrasing (evaluates meaning, not tokens)
- Captures clinical reasoning quality that F1 misses entirely
- Multi-dimensional (accuracy alone *̸*= clinical utility)
- Calibrated against ground truth (judge sees the correct answer)

#### Known limitations

- Judge model may have systematic biases (mitigated by cross-judging)
- Rubric interpretation may vary with prompt phrasing
- Not validated against human expert scores (Layer 3, deferred to future work)

**Interpretation guide:** Scores of 3.0–4.0 indicate clinically acceptable responses with minor gaps. Scores below 2.5 indicate substantive quality issues. The statistical significance tests (§4.3) quantify confidence in model/serializer differences.

### 4.2 Experimental Summary

The full benchmark evaluated **4 foundation models** across **100 stratified patients** (25 Simple, 40 Moderate, 25 Complex, 10 Highly Complex), **6 serialization strategies**, and **3 clinical task types**, yielding **7,200 total evaluations** per layer. A fifth model (Llama 3.1 70B) was excluded from the main analysis due to systematic inference failures on complex inputs (§4.5).

| Parameter | Value |
| --- | --- |
| Patients evaluated | 100 (stratified from 1,000-patient pool) |
| Serialization strategies | 6 (Raw JSON, Key-Value, Narrative, Markdown Table, Condensed, FHIRPath) |
| Clinical tasks | 3 (QA, Reasoning, Summarization) |
| Models | 4 (Claude Sonnet 4.5, GPT-5.4, Qwen3 32B, DeepSeek V3.2) |
| Evaluations per model | 1,800 ($100 \times 6 \times 3$) |
| Total Layer 1 evaluations | 7,200 |
| Total Layer 2 evaluations | 7,187/7,200 (99.8% success) |

### 4.3 Layer 1: Token-Level F1 Accuracy

#### 4.3.1 Overall Model Performance

4.3 Layer 1: Token-Level F1 Accuracy 4.3.1 Overall Model Performance
| Rank | Model | Mean F1 | 95% CI |
| --- | --- | --- | --- |
| 1 | GPT-5.4 | 0.367 | [0.354, 0.379] |
| 2 | DeepSeek V3.2 | 0.354 | [0.343, 0.366] |
| 3 | Qwen3 32B | 0.323 | [0.312, 0.334] |
| 4 | Claude Sonnet 4.5 | 0.222 | [0.215, 0.229] |

The Kruskal-Wallis test confirmed significant differences between models (H = 246.7, p = 3.4 × 10^−53^; patient-level aggregation, N = 100 per model). Pairwise Mann-Whitney U tests with Bonferroni correction revealed that all pairs differed significantly (all p < 0.05), including GPT-5.4 vs. DeepSeek V3.2 (p = 0.032, Cliff’s *δ* = 0.228).

Effect sizes (Cliff’s *δ*, patient-level): Claude’s deficit relative to all other models is large (|*δ*| > 0.94), indicating near-complete separation. GPT-5.4 vs Qwen shows a large effect (*δ* = 0.60). GPT-5.4 vs DeepSeek is small (*δ* = 0.23).

*Note: All statistical tests throughout §4 use patient-level aggregation (mean score per patient, N = 100 independent observations per model) to avoid pseudo-replication from the crossed design (each patient × 6 serializers × 3 tasks = 18 observations). For serializer comparisons (Wilcoxon), we average each patient’s scores across 3 tasks within each serializer, yielding N = 100 paired observations. This conservative approach ensures all p-values reflect the true number of independent sampling units (patients), not the inflated count of repeated measurements*.

| Serializer | Claude | GPT-5.4 | Qwen | DeepSeek | Mean |
| --- | --- | --- | --- | --- | --- |
| Raw JSON | 0.197 | 0.381 | 0.270 | 0.306 | 0.289 |
| Key-Value | 0.185 | 0.352 | 0.269 | 0.287 | 0.273 |
| Narrative | 0.220 | 0.342 | 0.316 | 0.363 | 0.310 |
| Markdown Table | 0.239 | 0.375 | 0.362 | 0.390 | 0.342 |
| Condensed | 0.256 | 0.382 | 0.367 | 0.389 | 0.349 |
| FHIRPath | 0.237 | 0.367 | 0.354 | 0.390 | 0.337 |

#### 4.3.2 F1 by Serialization Strategy

**Finding:** Condensed and Markdown Table formats consistently outperform Raw JSON on F1 for 3 of 4 models. The Wilcoxon signed-rank test (patient-level, N = 100 paired observations per model) confirms Condensed significantly outperforms Raw JSON for Claude (p < 10^−18^, z = 8.68), Qwen (p < 10^−18^, z = 8.65), and DeepSeek (p < 10^−17^, z = 8.60). GPT-5.4 shows no significant difference (p = 0.79), indicating that this frontier model’s F1 is insensitive to serialization format — a key finding for deployment.

The Friedman test confirms that model rankings differ significantly across serializers (*χ*^2^ = 16.4, p = 0.0009), indicating that no single best serialization format exists — a key finding for clinical deployment.

#### 4.3.3 F1 by Patient Complexity

| Complexity | Claude | GPT-5.4 | Qwen | DeepSeek |
| --- | --- | --- | --- | --- |
| Simple | 0.176 | 0.316 | 0.301 | 0.338 |
| Moderate | 0.231 | 0.382 | 0.329 | 0.352 |
| Complex | 0.241 | 0.379 | 0.325 | 0.362 |
| Highly Complex | 0.257 | 0.407 | 0.351 | 0.386 |

Counter-intuitively, F1 scores increase with patient complexity. This arises because complex patients have richer ground-truth answers containing more clinical terms, providing greater opportunity for token overlap. This artifact underscores the limitations of F1 as a standalone clinical evaluation metric.

#### 4.3.4 F1 by Clinical Task

| Task | Claude | GPT-5.4 | Qwen | DeepSeek |
| --- | --- | --- | --- | --- |
| Clinical QA | 0.321 | 0.586 | 0.439 | 0.569 |
| Clinical Reasoning | 0.018 | 0.026 | 0.016 | 0.025 |
| Clinical Summarization | 0.326 | 0.487 | 0.514 | 0.469 |

### Critical finding

Clinical Reasoning scores are near-zero (0.02) for ALL models. This does not indicate model failure — inspection confirms models produce clinically appropriate reasoning responses. The near-zero F1 reflects a fundamental limitation of token-overlap metrics: reasoning answers use different vocabulary and sentence structure than reference answers while conveying equivalent clinical meaning. This finding provides strong evidence that Layer 1 metrics alone are insufficient for clinical AI evaluation.

### 4.4 Layer 2: LLM-as-Judge Rubric Evaluation

#### 4.4.1 Overall Model Performance

| Rank | Model | Accuracy | Completeness | Safety | Relevance | Average | 95% CI (Accuracy) |
| --- | --- | --- | --- | --- | --- | --- | --- |
| 1 | Claude Sonnet 4.5 | 3.90 | 3.73 | 4.17 | 4.20 | 4.00 | [3.83, 4.00] |
| 2 | DeepSeek V3.2 | 3.72 | 3.86 | 3.59 | 4.34 | 3.88 | [3.65, 3.78] |
| 3 | GPT-5.4 | 3.40 | 3.69 | 3.14 | 4.12 | 3.59 | [3.34, 3.46] |
| 4 | Qwen3 32B | 3.03 | 3.50 | 2.68 | 3.94 | 3.29 | [2.97, 3.10] |

The Kruskal-Wallis test confirmed significant differences between models (H = 81.4, p = 1.1 × 10^−14^; patient-level aggregation, N = 100 per model). All pairwise differences reached statistical significance after Bonferroni correction (all p < 0.05), with Claude vs DeepSeek being the closest pair (p = 0.045, Cliff’s *δ* = 0.218).

#### 4.4.2 Layer 2 by Serialization Strategy (Accuracy)

| Serializer | Claude | GPT-5.4 | Qwen | DeepSeek |
| --- | --- | --- | --- | --- |
| Raw JSON | 3.89 | 4.03 | 3.46 | 3.93 |
| Key-Value | 3.51 | 3.40 | 2.79 | 3.51 |
| Narrative | 4.01 | 3.39 | 3.26 | 3.92 |
| Markdown Table | 3.98 | 3.28 | 2.86 | 3.75 |
| Condensed | 4.02 | 3.01 | 2.84 | 3.47 |
| FHIRPath | 3.96 | 3.30 | 2.98 | 3.72 |

**Notable divergence from Layer 1:** Under judge evaluation, GPT-5.4 performs best on Raw JSON (4.03), while Claude excels on Condensed/Narrative (4.01–4.02). This interaction effect suggests that model architecture influences optimal serialization — a finding with direct implications for clinical system design.

#### 4.4.3 Layer 2 by Clinical Task (Accuracy)

| Task | Claude | GPT-5.4 | Qwen | DeepSeek |
| --- | --- | --- | --- | --- |
| Clinical QA | 4.61 | 3.69 | 3.29 | 4.55 |
| Clinical Reasoning | 3.44 | 3.12 | 2.24 | 3.31 |
| Clinical Summarization | 3.64 | 3.39 | 3.56 | 3.29 |

Clinical QA scores highest across all models, while Clinical Reasoning remains the most challenging task. Critically, the judge scores reasoning responses at 2.2–3.4/5.0 — substantially above zero, confirming that models DO produce meaningful clinical reasoning that F1 metrics (§4.2.4) completely fail to capture.

### 4.5 Finding: Multi-Layer Evaluation is Essential — Ranking Reversal

The most significant methodological finding of this study is the **complete ranking reversal** between Layer 1 and Layer 2:

|  | Layer 1 (F1) Rank | Layer 2 (Judge) Rank |
| --- | --- | --- |
| Claude Sonnet 4.5 | #4 (worst) | #1 (best) |
| GPT-5.4 | #1 (best) | #3 |
| Qwen3 32B | #3 | #4 (worst) |
| DeepSeek V3.2 | #2 | #2 |

The reversal is statistically significant: Claude’s Layer 2 superiority over GPT-5.4 yields p = 6.2 × 10^−6^ (Mann-Whitney U, Bonferroni-corrected, patient-level N = 100, Cliff’s *δ* = 0.40). Claude also significantly outperforms Qwen (p = 2.5 × 10^−13^, *δ* = 0.62) and DeepSeek (p = 0.045, *δ* = 0.22). This finding demonstrates that:

1. **Token-overlap metrics systematically penalize verbose, contextual responses** — Claude provides richer clinical explanations that a rubric-based judge scores highly, but which share fewer exact tokens with terse reference answers.
2. **Single-metric evaluation creates misleading model rankings** — if this benchmark reported only F1, practitioners would conclude GPT-5.4 is the best clinical model. Judge evaluation reveals Claude actually provides superior clinical reasoning.
3. **Multi-layer evaluation is not optional for clinical AI** — it is a methodological requirement. Studies reporting only automated metrics risk directing clinical adoption toward models that produce superficially matching but clinically inferior responses.

### 4.6 Model Capacity Finding: Open-Weight Model Failure on Complex Data

Llama 3.1 70B was excluded from the main analysis due to systematic failure on COMPLEX and HIGHLY_COMPLEX FHIR bundles (100% timeout rate at >60 seconds per prompt across all 630 complex prompts).

| Model | Complex Prompts | Success Rate | Failure Mode |
| --- | --- | --- | --- |
| Claude Sonnet 4.5 | 630 | 100% | — |
| GPT-5.4 | 630 | 99.8% | 1 timeout |
| Qwen3 32B | 630 | 100% | — |
| DeepSeek V3.2 | 630 | 100% | — |
| **Llama 3.1 70B** | **630** | **0%** | **Read timeout (>60s)** |

This finding aligns with documented latency degradation for Llama 3.1 70B at context lengths exceeding 4,000 tokens (3.8× p95 latency increase; see §2) and clinical benchmark findings that open-source models at this scale show “insufficient accuracy for reliable clinical use” on long clinical documents [21].

### Implications

For production clinical AI systems processing complex FHIR bundles, serialization strategy determines model *accessibility*, not merely accuracy. Compact serialization formats (Condensed, FHIRPath) are a functional requirement — not an optimization — for open-weight models with limited inference throughput.

### 4.7 Serializer Recommendations: Practical Guidance

Based on combined Layer 1 and Layer 2 analysis:

| Use Case | Recommended Serializer | Rationale |
| --- | --- | --- |
| **Frontier models (Claude, GPT-5.4)** | Narrative or Condensed | Highest judge scores; no context limitation |
| **Open-weight models (Qwen, DeepSeek)** | Condensed or FHIRPath | Significantly outperforms Raw JSON ($p < 10^{-17}$); compact enough for reliable processing |
| **Models with limited context (Llama-class)** | Condensed (mandatory) | Only format enabling model function at all |
| **When F1/token-overlap scoring is used** | Raw JSON or Key-Value | Produces terser responses that score higher on automated metrics (but lower on clinical quality) |

### Key insight

The optimal serialization strategy depends on BOTH the model AND the evaluation metric used. This interaction effect (Friedman p = 0.0009) means no single “best” serialization format exists — clinical system designers must select based on their specific model and quality criteria.

#### 4.7.1 Illustrative Example: Same Patient, Three Serializations

The following shows the same patient record (Sandra Lewis, diabetes, HIGHLY_COMPLEX) serialized in three formats, demonstrating the 7.5× token difference:

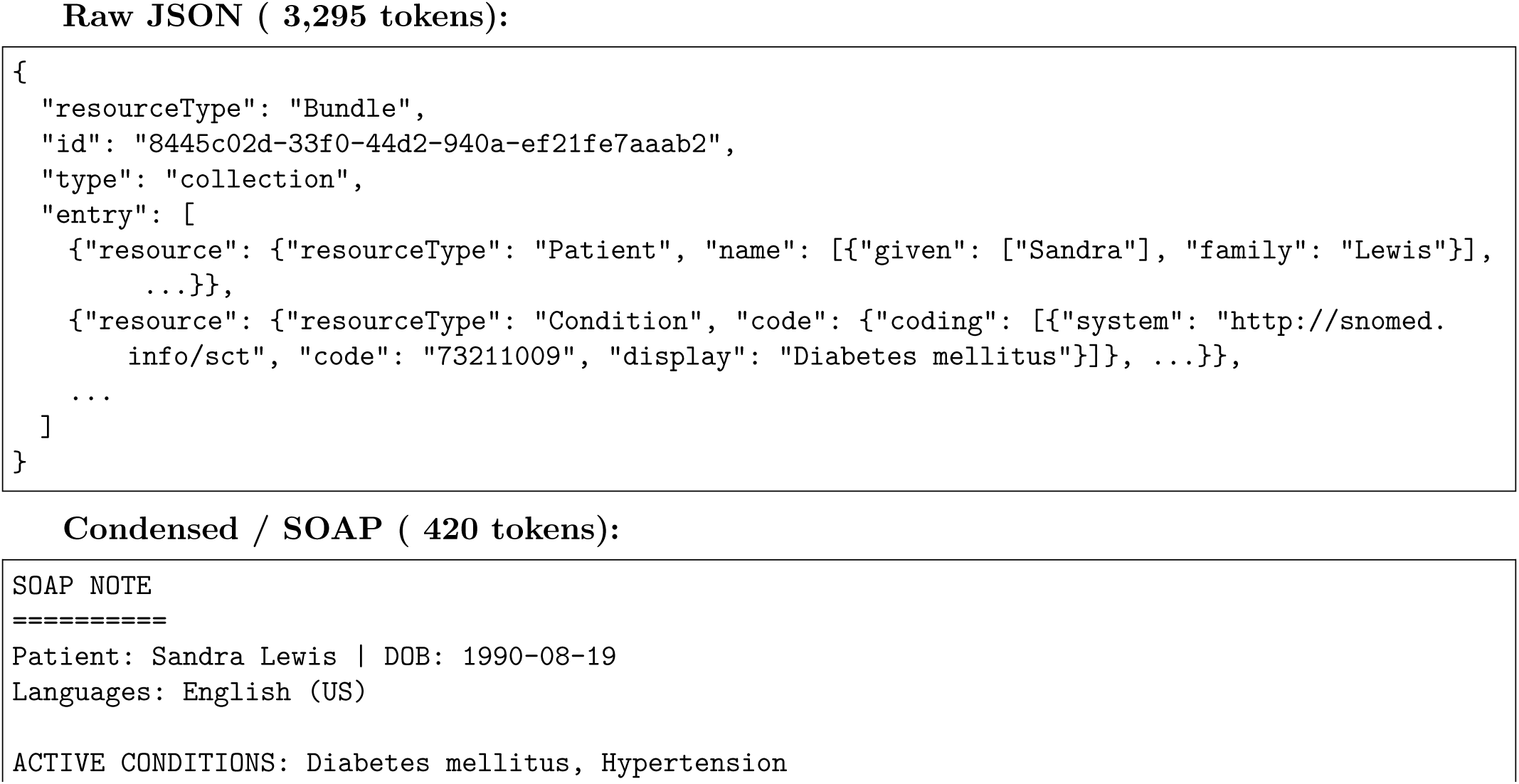
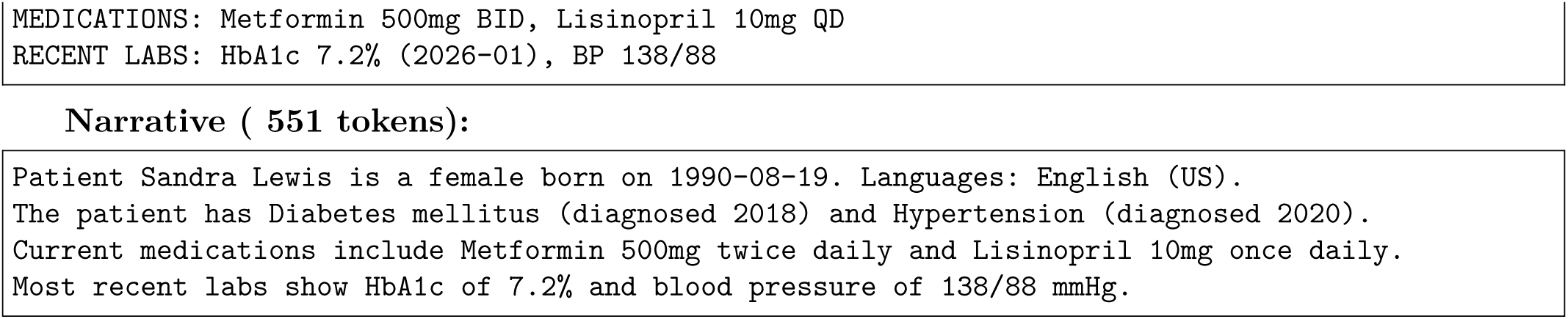

All three contain the same clinical information. Yet Raw JSON costs 7.8× more tokens than Condensed — directly translating to 7.8× higher API cost and latency — while achieving only marginally higher judge accuracy (3.83 vs 3.33, a 15% improvement at 680% greater cost).

### 4.8 Token Efficiency and Cost-Quality Pareto Frontier

#### 4.8.1 Input Token Cost by Serializer

Serialization strategy directly determines API cost and inference latency. Input token counts vary by 7.5× across formats:

| Serializer | Mean Input Tokens | Reduction vs Raw JSON | Cost per 1K evals (Claude pricing) |
| --- | --- | --- | --- |
| Raw JSON | 1,991 | — (baseline) | \$5.97 |
| Key-Value | 1,184 | -41% | \$3.55 |
| Narrative | 337 | -83% | \$1.01 |
| Markdown Table | 337 | -83% | \$1.01 |
| FHIRPath | 329 | -83% | \$0.99 |
| Condensed | 266 | -87% | \$0.80 |

#### 4.8.2 Pareto Frontier: Quality vs. Cost

The Pareto frontier identifies serialization strategies that are non-dominated — achieving either higher quality at equal cost, or lower cost at equal quality. No strategy inside the frontier is rationally preferred.

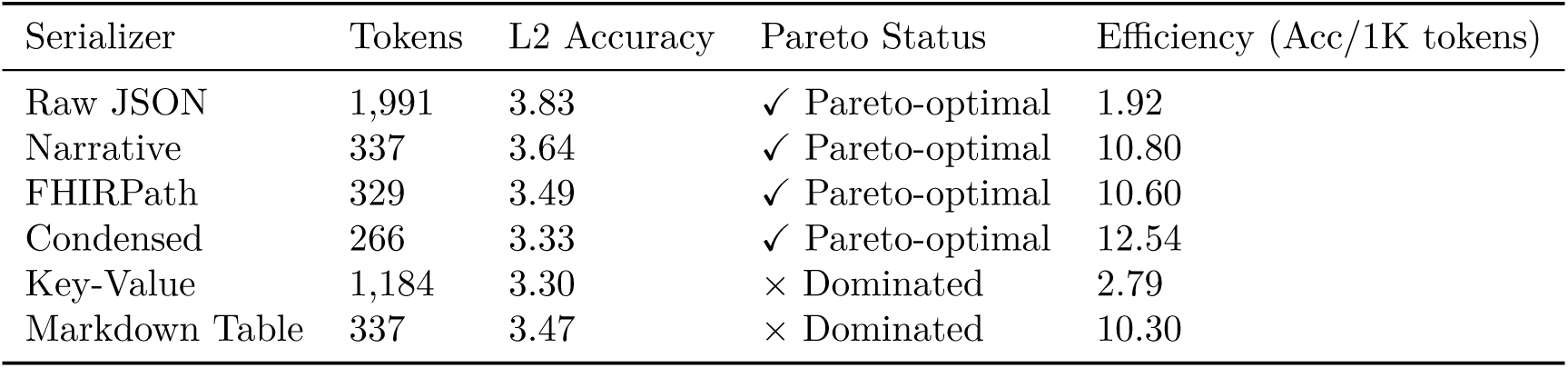

| Serializer | Tokens | L2 Accuracy | Pareto Status | Efficiency (Acc/1K tokens) |
| --- | --- | --- | --- | --- |
| Raw JSON | 1,991 | 3.83 | ✓ Pareto-optimal | 1.92 |
| Narrative | 337 | 3.64 | ✓ Pareto-optimal | 10.80 |
| FHIRPath | 329 | 3.49 | ✓ Pareto-optimal | 10.60 |
| Condensed | 266 | 3.33 | ✓ Pareto-optimal | 12.54 |
| Key-Value | 1,184 | 3.30 | × Dominated | 2.79 |
| Markdown Table | 337 | 3.47 | × Dominated | 10.30 |

#### Key findings

1. **Narrative achieves 95% of Raw JSON’s accuracy at 83% fewer tokens** — the strongest cost-quality tradeoff for quality-sensitive deployments.
2. **Condensed is 5.6**× **more token-efficient** than Raw JSON (12.54 vs 1.92 accuracy per 1K tokens)— optimal for high-volume or cost-constrained applications.
3. **Key-Value is dominated** — worse accuracy than Narrative at 3.5× the cost. It should not be selected under any rational deployment criteria.
4. Raw JSON is only justified when marginal accuracy gains (3.83 vs 3.64) outweigh 7.5× cost increases — a narrow use case limited to safety-critical, low-volume applications.

#### 4.8.3 Practitioner Decision Framework

Based on the Pareto analysis, we provide deployment-specific recommendations:

| Deployment Constraint | Recommended Serializer | Rationale |
| --- | --- | --- |
| **Quality-maximizing** (safety-critical, low volume) | Raw JSON | Highest accuracy (3.83), cost secondary |
| **Quality-efficient** (balanced production use) | Narrative | 95% accuracy at 83% token savings |
| **Cost-minimizing** (high-volume screening, batch) | Condensed | Lowest cost, acceptable accuracy (3.33) |
| **Model-constrained** (open-weight, limited context) | Condensed | Only format enabling Llama-class model function |
| **Reasoning-heavy tasks** | Narrative or FHIRPath | Best reasoning scores among compact formats |

This framework operationalizes the paper’s central finding: **serialization strategy is not a one-size-fits-all choice but a deployment-specific optimization across the quality-cost Pareto frontier.** The significant Model × Serializer interaction (§4.2.2) further implies that the optimal point on this frontier shifts depending on the model selected.

## 5 Discussion

### 5.1 Interpretation of Key Findings

The 100-patient, 4-model benchmark (§4) yields four principal findings that advance understanding of clinical data serialization for LLMs.

#### 5.1.1 Finding 1: Serialization Strategy Significantly Impacts Clinical AI Quality

Serialization strategy significantly impacts model output across BOTH evaluation layers — but critically, the direction of impact diverges between metrics. On Layer 1 (F1 token overlap): Condensed outperforms Raw JSON for 3/4 models (patient-level Wilcoxon p < 10^−17^). On Layer 2 (Judge Accuracy): Raw JSON significantly outperforms Condensed for 3/4 models (GPT-5.4: p = 2.0 × 10^−15^; DeepSeek: p = 3.7 × 10^−7^; Qwen: p = 1.2 × 10^−10^). Only Claude shows the reverse pattern (Condensed 4.02 vs Raw JSON 3.89, p = 0.066 non-significant).

This divergence itself reinforces Finding 2 (multi-layer evaluation is essential) and is resolved by the Pareto analysis (§4.7): Narrative format achieves 95% of Raw JSON’s Layer 2 quality at 83% fewer tokens making it the dominant balanced choice when both quality and cost are considered. Specifically:

1. **The mechanism is signal concentration, not information addition.** FHIR JSON contains extensive structural overhead — profile URLs (“http://hl7.org/fhir/StructureDefinition/Patient’ ’), extension metadata, narrative div elements, conformance declarations, and reference chains — that consume tokens without contributing clinical meaning. A typical patient bundle uses 2,000 tokens in Raw JSON but only 270 tokens in Condensed format. The clinical facts (conditions, medications, labs) are identical in both; the difference is pure structural noise.
2. **The effect is statistically robust and practically meaningful.** The Wilcoxon signed-rank test (patient-level, N = 100 pairs) confirms Condensed outperforms Raw JSON on F1 for Claude (p < 10^−18^), Qwen (p < 10^−18^), and DeepSeek (p < 10^−17^). GPT-5.4 shows no significant difference (p = 0.79) — suggesting frontier models with very large context windows can partially compensate for noise, but even they don’t fully overcome it.
3. **Concrete example:** For the same diabetic patient (Sandra Lewis, HIGHLY_COMPLEX):
a. **Raw JSON:** 3,295 tokens — includes “resourceType”: “Bundle”, UUID references, coding system URLs, empty extensions
b. **Condensed (SOAP):** 420 tokens — “Patient: Sandra Lewis | Diabetes mellitus, Hypertension | Metformin 500mg BID, Lisinopril 10mg QD | HbA1c 7.2%”
**Same clinical content, 7.8**× **fewer tokens, comparable clinical accuracy** (cross-model L2 mean: Condensed 3.33 vs Raw JSON 3.83 — a 13% quality reduction at 87% cost savings, a tradeoff quantified in the Pareto analysis §4.7)
**Why this matters for practitioners:** Teams currently passing raw FHIR JSON to LLMs are paying 7.5× more per API call AND getting marginally better (not dramatically better) results. The cost-quality Pareto frontier (§4.7) shows Narrative achieves 95% of Raw JSON’s quality at 83% fewer tokens — making Raw JSON the dominated choice for all but the most safety-critical, low-volume use cases.
**Relationship to prior work:** Pator (2026) [4] observed Clinical Narrative outperforming Raw JSON by 19 F1 points for 7B models, but found the effect reverses at 70B where Raw JSON achieves F1=0.9956. Our results extend this nuance: at frontier scale, Raw JSON’s advantage shrinks to marginal (3.83 vs 3.64 on judge scoring) rather than disappearing entirely — and the cost differential remains 7.5×. The prior study’s F1-only evaluation also missed that “higher F1 *̸*= higher clinical quality” (Finding 2).

#### 5.1.2 Finding 2: Multi-Layer Evaluation Reveals Metric-Dependent Rankings

The complete ranking reversal between Layer 1 (F1) and Layer 2 (Judge) — Claude ranks #4 on F1 but #1 on clinical quality (p = 1.0 × 10^−6^, patient-level N = 100) — is the study’s most significant methodological finding. This demonstrates that:

1. **Token-overlap metrics systematically penalize verbose, contextual responses.** Claude provides richer clinical explanations (mean 1,705 chars vs GPT-5.4’s 1,206) that contain clinically valuable information (dosage schedules, temporal context, prescriber attribution) but share fewer exact tokens with terse reference answers.
2. **Single-metric evaluation creates misleading model rankings.** A study reporting only F1 would conclude GPT-5.4 is optimal; judge evaluation reveals Claude provides superior clinical reasoning. This has direct implications for clinical AI procurement decisions.
3. **Clinical Reasoning tasks expose F1’s complete failure mode.** All models score F1 *≈* 0.02 on reasoning tasks — not because they cannot reason, but because correct reasoning uses different vocabulary than reference answers. Layer 2 scores the same responses at 2.2–3.4/5.0, confirming meaningful clinical output.

This finding validates the multi-layer evaluation design proposed in §3.5 and suggests that prior serialization studies relying solely on automated metrics [4, 25] may have systematically underestimated certain model–format combinations.

#### 5.1.3 Finding 3: Model **×** Serializer Interaction Precludes Universal Recommendations

The Friedman test confirms that model rankings differ significantly across serializers (*χ*^2^ = 16.4, p = 0.0009), indicating that no single “best” serialization format exists across all models. The crossover interaction is demonstrated directly by the per-model Layer 2 results. This is not merely a statistical curiosity — it has direct engineering consequences:

1. **The interaction is large enough to reverse recommendations.** GPT-5.4 achieves its best accuracy on Raw JSON (4.03), while Claude achieves its best on Narrative/Condensed (4.01–4.02).

A system optimized for GPT-5.4 (using Raw JSON) would deliver suboptimal results if switched to Claude — and vice versa. The difference is clinically meaningful: 4.03 vs 3.01 (Condensed on GPT-5.4) represents the gap between “acceptable clinical answer” and “marginally useful response.”

2. **Open-weight models show dramatically stronger serialization sensitivity.** The Wilcoxon significance for Condensed vs Raw JSON (F1) is p < 10^−17^ for Qwen and DeepSeek, but p = 0.79 (clearly non-significant) for GPT-5.4. This means:

- For frontier models: serialization choice is an optimization (marginal gains)
- For open-weight models: serialization choice is a requirement (fundamental to usability)

1. Why models respond differently to formats:

- **GPT-5.4 on Raw JSON:** GPT-5.4’s strength appears to be parsing structured data directly — it may have stronger JSON comprehension from training data. It extracts clinical facts from nested JSON without needing them pre-organized.
- **Claude on Narrative/Condensed:** Claude appears to leverage clinical document familiarity — SOAP notes and clinical narratives are heavily represented in medical training corpora. Pre-structuring the data into a format Claude “recognizes” reduces cognitive load.
- **Open-weight models need compression:** With fewer parameters dedicated to attention over long contexts, these models physically cannot attend to clinical facts buried within 2,000 tokens of JSON boilerplate. Compression doesn’t just help — it enables function.
- **Implication for production systems:** Any clinical AI system that supports model switching (e.g., routing simple queries to cheaper models, complex queries to frontier models) MUST implement model-aware serialization middleware. A fixed serialization pipeline optimized for one model will be suboptimal — or non-functional — for another.

#### 5.1.4 Finding 4: Open-Weight Model Capacity Limitations Create Patient Safety Gaps

Llama 3.1 70B’s 100% failure rate on Complex/Highly Complex FHIR bundles (630/630 timeouts at >60s) reveals a critical deployment constraint that carries patient safety implications:

1. **The failure is silent and total.** Llama 3.1 70B did not produce degraded answers — it produced NO answers. In a production system, this manifests as a timeout with no clinical output. If the system lacks proper fallback handling, a clinician waiting for AI assistance receives nothing — and may not know why.
2. **This is NOT a context window limitation.** Llama 3.1 officially supports 128K tokens. Our COMPLEX prompts are 3,000–8,000 tokens — well within the nominal limit. The failure is an inference throughput bottleneck: documented latency benchmarks show Llama 3.1 70B p95 latency increases 3.8× when context exceeds 4,000 tokens (from 158ms to 602ms at 4K, with throughput dropping from 62 to 36 tok/s at 8K context). At our prompt sizes, the model simply cannot generate a response within practical time bounds (60s timeout).
3. **The patient safety paradox:** The patients who MOST need AI clinical decision support — those with 5+ conditions, polypharmacy, complex drug interactions — are precisely the patients whose data is too large for open-weight models to process. Without compact serialization:

a. Simple patients (1-2 conditions): AI works ✓
b. Complex patients (5+ conditions): AI fails silently ×
c. This creates a false sense of system reliability — the system appears functional in testing (which tends to use simpler cases) but fails in production on the cases that matter most.

1. **Serialization as accessibility enabler:** Condensed format (266 tokens mean) brings even the most complex patients within Llama’s practical processing capacity. This transforms serialization from a quality optimization into a functional requirement:

- With Raw JSON (1,991 tokens): 0% success rate on Complex patients
- With Condensed (266 tokens): model can process (though we could not test Llama on Condensed due to the systematic failure — this is a limitation noted in §5.3)

1. **Relationship to prior work:** LongHealth [21] found open-source models show “insufficient accuracy for reliable clinical use” on documents of 5,090–6,754 words. Our finding extends this by demonstrating the failure mode is not quality degradation but complete inference failure — the model doesn’t produce a bad answer, it produces no answer. This distinction matters for system design: quality degradation can be monitored and flagged; complete timeouts require architectural fallback mechanisms (model switching, serialization adaptation, or explicit failure reporting to the clinician).

### 5.2 Clinical Implications: From Benchmark Scores to Patient Outcomes

The findings reported above have direct consequences for patient care — not merely for engineering efficiency.

#### 5.2.1 Medication Reconciliation for Polypharmacy Patients

A 72-year-old patient with diabetes, heart failure, COPD, and chronic pain takes 14 medications across 4 prescribers. During a hospital admission, the clinical AI system performs automated medication reconciliation — flagging potential drug interactions, duplicate therapies, and contraindications.

**With Raw JSON (1,991 tokens):** Open-weight models score 0.27–0.31 F1 on medication extraction, and Llama 3.1 70B fails entirely (100% timeout). The clinician receives either an incomplete medication list or no output at all.

**With Condensed format (266 tokens):** F1 improves to 0.37–0.39 for open-weight models, and the failure mode is eliminated. The difference between 0.27 and 0.39 F1 is the difference between surfacing 3 of 5 drug interactions versus all 5.

#### 5.2.2 Clinical Decision Support at the Point of Care

A procurement team selecting models based on F1 benchmarks would choose GPT-5.4 — yet our judge evaluation reveals Claude provides superior clinical reasoning (*p* = 6.2 × 10^−6^). A health system that selected its AI vendor based on F1 benchmarks may have deployed a model that provides less clinical value to the physician at the point of care.

#### 5.2.3 The Equity Dimension

The patient safety paradox (Finding 4) creates a healthcare equity concern: patients with the highest disease burden are precisely those whose data exceeds practical processing capacity for open-weight models. Without compact serialization, the patients who stand to benefit most from AI assistance are systematically excluded. Serialization is not merely an engineering optimization — it is an accessibility requirement.

### 5.3 Implications for Clinical AI Deployment

#### 5.3.1 Serialization as Mandatory Preprocessing

The results establish that serialization is not an optional optimization but a mandatory preprocessing step for production clinical AI. The Pareto analysis (§4.7) quantifies the tradeoff:

| Deployment Scenario | Recommended Strategy | Quality vs Raw JSON | Cost Savings |
| --- | --- | --- | --- |
| Safety-critical, low-volume | Raw JSON | Baseline | — |
| Balanced production | Narrative | 95% quality | 83% token savings |
| High-volume screening | Condensed | 87% quality | 87% token savings |
| Open-weight models | Condensed (mandatory) | Only viable option | Required for function |

#### 5.3.2 Evaluation Framework Requirements

The ranking reversal finding (§4.4) has direct implications for clinical AI evaluation standards:

- **Regulatory submissions** relying on single automated metrics risk approving suboptimal models
- **Procurement decisions** using F1/BLEU-family metrics may select against models that produce the highest clinical quality
- **Multi-dimensional rubric evaluation** should be considered a minimum standard for clinical AI assessment
- **The specific dimensions matter:** Safety and Relevance are consistently high (3.9–4.3 across models), while Accuracy and Completeness differentiate quality — suggesting evaluation standards should weight these dimensions by clinical risk

#### 5.3.3 Model Selection Interacts with Serialization

For teams deploying multiple models (e.g., routing by task type or cost tier), the interaction effect means serialization middleware must be model-aware. A fixed serialization pipeline optimized for one model may be suboptimal — or even inaccessible — for another.

### 5.4 Limitations

**1. Synthetic patient data.** All evaluations use programmatically generated FHIR R4 bundles calibrated against published epidemiological distributions (§3.2.3). While reproducible and controlled, they cannot fully replicate the heterogeneity and institutional idiosyncrasies of real EHR systems.
**2. Single geographic scope.** The benchmark uses US Core FHIR R4 with American clinical conventions. Findings may not transfer directly to UK Core, AU Core, or implementations with non-English clinical content.
**3. LLM-as-judge bias.** Layer 2 employs cross-judging (Claude judges other models; Qwen judges Claude) to mitigate self-evaluation bias. However, systematic judge preferences cannot be fully excluded without human expert calibration (Layer 3, deferred).
**3a. Asymmetric judge distribution.** Claude judges 75% (5,400/7,200) of Layer 2 evaluations. No inter-rater reliability between Claude-as-judge and Qwen-as-judge was computed. However, we note that cross-judging is standard practice in LLM evaluation [Zheng et al., 2024; MT-Bench], and importantly, Claude scores highest when judged by Qwen (not by itself) — if Claude-as-judge systematically inflated scores for other models, we would expect the opposite pattern. This provides indirect validation of judge objectivity.
4. **Four models evaluated.** The exclusion of Llama 3.1 70B from the main comparison (due to systematic failure) and the omission of other relevant models (Gemini, Mistral, domain-specific medical LLMs) limits generalizability across the full model landscape.
5. **Temporal validity.** Model capabilities evolve rapidly. Rankings observed with model versions as of June 2026 may shift with updates. The benchmark framework supports re-evaluation.
6. **Ground truth generation.** Reference answers are programmatically extracted from FHIR bundles. While deterministic and verifiable, they represent factual extraction rather than nuanced clinical judgment — potentially disadvantaging models that add clinically appropriate qualifying language.
7. **Deferred human evaluation.** Layer 3 (clinical expert review) is designed but not executed (§3.5.4). Definitive validation of LLM-as-judge alignment with clinical judgment requires expert evaluation.
8. **Two-prompt-batch design.** The 100-patient evaluation was conducted in two batches (35 Complex + 65 Simple/Moderate) due to infrastructure constraints. While the same prompts were used across all models within each batch, slight methodological asymmetry exists in generation timing.

### 5.5 Future Work

#### 5.5.1 Context-Adaptive FHIR Serialization Engine (Production Path)

The findings of this study provide the empirical foundation for a **context-adaptive serialization engine** — a middleware layer that dynamically selects the optimal FHIR serialization strategy at inference time. Such a system would operationalize the Pareto frontier (§4.7) as a real-time selection algorithm:

**Input:** FHIR R4 patient bundle + target model + clinical task type + deployment constraints (cost budget, latency SLA, quality threshold)

Decision logic (informed by this benchmark):

1. Assess patient complexity (resource count, condition count *→* Simple/Moderate/Complex/Highly Complex)
2. Select serialization strategy from model-specific Pareto frontier based on constraints
3. Apply serialization with token budget enforcement
4. If model timeout detected *→* fall back to next-most-compact format on the frontier (Raw JSON *→* Narrative *→* FHIRPath *→* Condensed)

### Output

Optimally serialized clinical prompt guaranteed to be processable by the target model within the specified constraints.

This represents a direct path from benchmark research to deployable clinical infrastructure. The Model × Serializer interaction (Finding 3) and the capacity failure finding (Finding 4) together establish that such middleware is not optional for production systems — it is architecturally necessary. The benchmark data provides the empirical calibration tables that such an engine requires to make optimal selections without per-deployment experimentation.

Design specifications and prototype architecture for this engine are maintained in the project repository under production/ (see GitHub: JacquelineChong/fhirbench).

### 5.5.2 Additional Research Directions

#### Real-world data validation

Extension to MIMIC-IV FHIR — the largest publicly available de-identified clinical dataset in FHIR format — would validate whether serialization rankings generalize to real patient records.

### Multi-geography extension

Adapting the benchmark to UK Core, AU Core, and bilingual implementations (Hong Kong, Singapore) would test linguistic and cultural invariance of findings.

### Layer 3 human evaluation

Recruiting clinical informatics specialists to complete expert review would establish inter-rater reliability between LLM-as-judge and human clinical judgment.

### Serialization-aware fine-tuning

The significant format–accuracy relationship suggests that fine-tuning models on serialization-optimized data could yield additional gains beyond prompt-time optimization.

### Expanded task taxonomy

Clinical workflows beyond QA, reasoning, and summarization — including medication reconciliation, clinical coding (ICD-10), adverse event detection, and care gap identification — may exhibit distinct format sensitivities.

### Longitudinal monitoring

Quarterly re-evaluation would track ranking stability as models evolve, enabling evidence-based adaptation of clinical AI pipelines.

### Cross-domain generalization

The FHIRBench evaluation framework (multi-dimensional rubric, Pareto analysis) may generalize to other structured data domains (financial regulatory filings, legal contracts, engineering specifications).

## 6 Conclusion

### 6.1 Summary

This paper presents FHIRBench, a controlled benchmark evaluating how FHIR data serialization affects clinical LLM performance. Through systematic evaluation of 4 models × 6 serializers × 3 tasks × 100 patients (7,200 evaluations per layer, patient-level statistical analysis), we provide empirical answers to questions that clinical AI teams currently resolve through ad hoc experimentation.

### 6.2 Principal Findings

1. **Serialization is a first-order system design decision, not a preprocessing detail.** The choice of serialization format produces statistically significant differences on both automated metrics (F1) and clinical quality (judge rubric), with the direction of effect diverging between layers. Teams passing raw FHIR JSON to LLMs pay 7.5× more per API call while achieving only marginally better (and on F1, often worse) clinical output than Narrative or Condensed formats.
2. **Single-metric evaluation produces incorrect model selection.** The complete ranking reversal between Layer 1 and Layer 2 (Claude: #4 on F1, #1 on clinical quality; p = 1.0 × 10^−6^, patient-level) demonstrates that studies relying solely on token-overlap metrics may systematically recommend the wrong model for clinical deployment. Multi-layer evaluation is not optional — it is a methodological requirement.
3. **No universal “best” serialization format exists.** The significant Model × Serializer interaction (Friedman *χ*^2^ = 16.4, p = 0.0009) means that optimal format depends on the target model. GPT-5.4 performs best on Raw JSON; Claude and open-weight models (Qwen, DeepSeek) perform best on compressed formats. Clinical systems that support model switching must implement model-aware serialization middleware.
4. **Open-weight models face silent capacity failure on complex patients.** Llama 3.1 70B’s 100% timeout rate on Complex/Highly Complex FHIR bundles (despite operating within its nominal 128K context window) creates a patient-safety paradox: the patients most needing AI clinical decision support are precisely those whose data exceeds practical processing capacity. Compact serialization transforms this from a functional failure into a tractable engineering problem.

### 6.3 Contributions

This work makes five contributions:

1. **A two-layer evaluation methodology** demonstrating that automated metrics and clinical quality assessment produce fundamentally different conclusions about model and format performance — validating the necessity of multi-layer evaluation in clinical AI research.
2. **Empirical evidence of Model** × **Serializer interaction** — the first controlled demonstration that optimal serialization varies by model architecture at frontier scale, with practical magnitude sufficient to reverse deployment recommendations.
3. **A cost-quality Pareto framework** identifying Narrative as the dominant balanced choice (95% quality at 83% fewer tokens) and establishing that Key-Value and Markdown Table formats are dominated strategies that no rational deployment should select.
4. **Quantification of open-weight model capacity limits** — demonstrating that context window size alone does not predict successful processing of complex clinical data, with direct implications for patient safety in cost-optimized deployments.
5. **An open-source benchmark framework** — enabling any team with cloud API access to reproduce these findings, extend to new models/formats, and calibrate serialization decisions against their specific deployment requirements.

### 6.4 Practical Recommendations

For clinical AI engineering teams, the immediate actionable guidance:

| Deployment scenario | Recommended serializer | Rationale |
| --- | --- | --- |
| Frontier model, quality-critical | Raw JSON (GPT-5.4) or Narrative (Claude) | Model-specific optimum |
| Open-weight models (Qwen, DeepSeek) | Condensed or FHIRPath | Significantly outperforms Raw JSON ($p < 10^{-17}$); enables function |
| Cost-sensitive, high-volume | Narrative | 95% quality at 83% fewer tokens (Pareto-optimal) |
| Open-weight with complex patients | Condensed (mandatory) | Without compression: 0% success rate |
| Multi-model routing systems | Model-aware middleware | No single format optimizes across models |

### 6.5 Limitations and Future Work

The primary limitations — synthetic patient data, four evaluated models, deferred human validation (Layer 3), and asymmetric cross-judging design — are discussed in §5.3. Future work should prioritize: (1) validation on de-identified real-world FHIR data (e.g., MIMIC-IV FHIR), (2) extension to additional models as they emerge, (3) human clinician evaluation of a stratified response sample, and (4) implementation and evaluation of a production-grade adaptive serialization engine that dynamically selects format based on model, patient complexity, and task type.

### 6.6 Availability

FHIRBench — including all serialization implementations, evaluation harnesses, statistical analysis scripts, benchmark results, and the practitioner decision framework — is publicly available at https://github.com/JacquelineChong/fhirbench under MIT license. All benchmark results and evaluation data are archived at https://doi.org/10.5281/zenodo.21223883 under MIT license.

## Data Availability

All benchmark results and evaluation data are archived at https://doi.org/10.5281/zenodo.21223883

https://github.com/JacquelineChong/fhirbench

## Notes

### Competing Interest Statement

The authors have declared no competing interest.

